# Psychological impact of infectious disease outbreaks on pregnant women: Rapid evidence review

**DOI:** 10.1101/2020.04.16.20068031

**Authors:** Samantha K. Brooks, Dale Weston, Neil Greenberg

**Author notes:** Corresponding author: Dr Samantha K. Brooks, Department of Psychological Medicine, King’s College London, Weston Education Centre, London, SE5 9RJ, UK.

## Abstract

Infectious disease outbreaks can be distressing for everyone, especially so for those deemed to be particularly vulnerable, such as pregnant women who have been named a high-risk group in the current COVID-19 pandemic. This rapid review aimed to summarise existing literature on the psychological impact of infectious disease outbreaks on women who were pregnant at the time of the outbreak. In April 2020 five databases were searched for relevant literature and main findings were extracted. Thirteen papers were included in the review. The following themes were identified: negative emotional states; living with uncertainty; concerns about infection; concerns about and uptake of prophylaxis or treatment; disrupted routines; non-pharmaceutical protective behaviours; social support; demands from others; financial and occupational concerns; disrupted expectations of birth, prenatal care and postnatal care, and; sources of information. Results showed that pregnant women have unique needs during infectious disease outbreaks and could benefit from: up-to-date, consistent information and guidance; appropriate support and advice from healthcare professionals, particularly with regards to the risks and benefits of prophylaxis and treatment; virtual support groups, and; designating locations or staff specifically for pregnant women.

## Introduction

Infectious disease outbreaks pose a major health risk across the globe. The outbreak of coronavirus COVID-19 was declared a pandemic by the World Health Organization on March 11^th^ 2020 and has – as of April 3^rd^ 2020 - seen over a million confirmed cases and over 58,000 related deaths worldwide (Rawlinson, 2020). Such outbreaks are understandably distressing for many people; there is a wealth of research to suggest a substantial negative psychological impact of pandemics (Gardner & Moallef, 2015; Perrin et al., 2009).

There is historical evidence of pregnant women being classified as a high-risk group during pandemics; for example, pregnancy was associated with poor clinical outcomes and high mortality rates during the H1N1 ‘swine flu’ pandemic (Mosby et al., 2011) and the severe acute respiratory syndrome (SARS) pandemic (Lam et al., 2004; World Health Organization, 2010). Indeed, during the COVID-19 outbreak questions have also been raised about the particular risks for pregnant women and their unborn babies (Schwartz & Graham, 2020). A review of the limited data available so far (Dashraath et al., 2020) suggests that COVID-19 outcomes for mothers are more promising than for previous outbreaks. They report relatively low foetal complications associated with infection and report that 46/48 neonates born to COVID-19 infected mothers have not tested positive. They note that the majority of these women acquired COVID-19 in the third trimester; there is currently no data on outcomes for infections acquired in early pregnancy. Despite this promising early data, more research is needed to ascertain the true risk of COVID-19 for pregnant women and their unborn babies. As a result of this uncertainty, on March 16^th^ 2020 the UK government announced that pregnant women were considered a ‘vulnerable group’ who should be shielded from the virus (Jarvis, 2020; Public Health England, 2020).

Although there is a considerable body of literature on the clinical course and outcomes of being diagnosed with an infectious disease during pregnancy, comparatively little attention has been paid to the potential psychological impact such outbreaks can have on pregnant women, regardless of their infection status. However, it is likely that pregnant women may experience particularly high levels of distress during an infectious disease outbreak for several reasons. Firstly, they may have serious concerns about their own health, given the physiological changes that occur during pregnancy which may make them more susceptible to, or more severely affected by, infectious diseases (Jamieson et al., 2006a). They may also have concerns about the health of their unborn babies. Additionally, public health recommendations during infectious disease outbreaks often involve the use of novel vaccines and medications for prophylaxis and treatment, meaning that pregnant women are faced with the dilemma of either not complying with recommendations - thus putting themselves at risk of infection - or using novel medical treatments with little evidence available on the impact they might have on an unborn baby. Pregnant women may also experience distress due to the disruption of routine prenatal care and delivery: for example, women who gave birth in Hong Kong during the SARS outbreak were discharged as soon as possible after delivery and all prenatal services considered non-essential were suspended (Jamieson et al., 2006b), and hospitals in Taiwan saw a significant reduction in length of stay after birth (Lee et al., 2005). Similarly, during the H1N1 pandemic, the World Health Organization (2010) provided guidelines to programme managers and clinicians suggesting that both antenatal clinic visits and time spent in hospital by mother and baby immediately post-birth be reduced to the minimum required.

The COVID-19 pandemic may also lead to pregnant women becoming concerned because of social distancing and self-isolation recommended by governments worldwide. Rasmussen et al. (2008) point out that, even if medical appointments are reduced to the minimum possible, pregnant women continue to require both outpatient prenatal care and inpatient delivery services during a pandemic; in the context of COVID-19 this necessitates pregnant women leaving the safety of self-isolation. Additionally, research suggests that lack of control, or powerlessness over decisions relating to childbirth, can be traumatic (Patterson et al., 2018) raising concerns about how women giving birth during the COVID-19 pandemic will cope with decisions over the birthing process being out of their control. Furthermore, the COVID-19-related restrictions on hospital visitation procedures may be distressing for pregnant women; in many countries (such as the UK) women are being requested to attend all prenatal appointments alone (Oppenheim, 2020), and in some countries (including Poland and China) are even being required to give birth alone (Kahn & Cristoferi, 2020; Stevenson, 2020). However, familial support during the birthing process (including labour, delivery, and post-partum) is considered essential for women’s wellbeing (American College of Obstetricians and Gynecologists, 2017) and the lack of such support may be difficult to cope with.

Given the relative dearth of research on the subject, the current review represents the first attempt to systematically examine literature on the psychological impact of previous infectious disease outbreaks on pregnant women. Specifically, the two main aims of the review were to consider i) what are the psychological outcomes associated with being pregnant during an infectious disease outbreak and ii) what factors are associated with these psychological outcomes, in order to gain an idea of how to reduce any negative psychological outcomes for pregnant women.

## Method

This research was carried out as a rapid evidence review in response to the COVID-19 outbreak of 2019-2020. Rapid reviews follow the general guidelines for traditional systematic reviews but are simplified (e.g. not searching grey literature; not conducting quality appraisal of the included studies; not translating foreign-language papers) in order to produce evidence rapidly. Rapid reviews are recommended during circumstances where policy-makers urgently need evidence synthesis as quickly as possible in order to inform public health guidelines (World Health Organization, 2017).

### Search strategy

The following databases were searched: Medline, PsycInfo, Embase, Global Health and Web of Science. All were searched from inception to the date of the searches (1 April 2020).

Search terms were:

1. pregnant OR pregnanc*
2. psychological OR mental health OR trauma OR stress OR distress OR anxiety OR wellbeing OR well-being OR panic OR depress*
3. pandemic* OR disease outbreak* OR SARS OR severe acute respiratory syndrome OR swine flu OR H1N1 OR avian influenza OR bird flu OR H5N1 OR Ebola OR MERS OR Middle East respiratory syndrome OR Zika OR coronavirus OR COVID-19
4. 1 AND 2 AND 3

### Selection criteria

To be included in the review, studies had to: i) report primary data; ii) be published in peer-reviewed journals; iii) be written in English; and iv) report on any psychological effects of emerging infectious disease outbreaks on women who were pregnant at the time of the outbreak. Infectious disease outbreaks included SARS, MERS and Ebola. Zika was also considered a relevant outbreak for review because, despite the spread of the infection being different (i.e. primarily through mosquito bites rather than spread from person to person) this outbreak had a particular impact on pregnant women as it was associated with birth defects (Chakhtoura et al., 2019). The review focused on emerging infectious diseases (that is, one that appears for the first time in a population or rapidly increases in incidence or geographic range; World Health Organization, 2014), therefore studies looking at seasonal influenza were excluded.

### Screening

The authors ran the search strategy and downloaded citations from all databases to EndNote version X9 (Thomson Reuters, New York, United States) where duplicates were automatically removed. Titles and then abstracts were screened for their relevance to the selection criteria by one author (SKB). Full-texts of all papers remaining after abstract screening were downloaded and assessed to decide whether they met all inclusion criteria. Reference lists of all included papers were hand-searched.

### Data extraction and synthesis

Spreadsheets were created in order to systematically extract the following data from papers: authors, year of publication, country of study, design, infectious disease outbreak studied, number of participants, socio-demographic information on participants, measures used, and key results. The results of the studies were synthesised by one author (SKB) using thematic analysis to code the data and organise into themes (Braun & Clarke, 2006).

## Results

A total of 1694 citations were found in the initial search and 560 duplicates removed. 999 papers were excluded based on title, 115 excluded based on abstract, and nine excluded after assessing full-texts. Two additional papers were found via hand-searching, leaving a total of thirteen papers included in the review. Studies were international, including three from China (Dodgson et al., 2010; Lee et al., 2006; Ng et al., 2013; one with participants from Brazil, Puerto Rico and the USA (Linde & Siqueira, 2018); one with participants from both Scotland and Australia (Lohm et al., 2014); three from the USA (Lyerly et al., 2012; Lynch et al., 2012; Steelfisher et al., 2011); one from Brazil (Meireles et al., 2017); one from Turkey (Ozer et al., 2010); one from Canada (Sakaguchi et al., 2011); one from Japan (Sasaki et al., 2013); and one from both Scotland and Poland (Sim et al., 2011). Three focused on the SARS outbreak, eight papers on H1N1 and two papers on the Zika virus.

An overview of study characteristics is presented in Table I, while a detailed summary of the thematic analysis is presented in Table II. It should be noted that there is some overlap between themes; specifically, ‘negative emotional states’ in general emerged as a theme, but many papers also referred to negative emotions due to specific factors, which have also been classified as themes (for example, ‘concern about risk of infection’ is a negative emotional state, but also a theme in itself).

**Table I.**
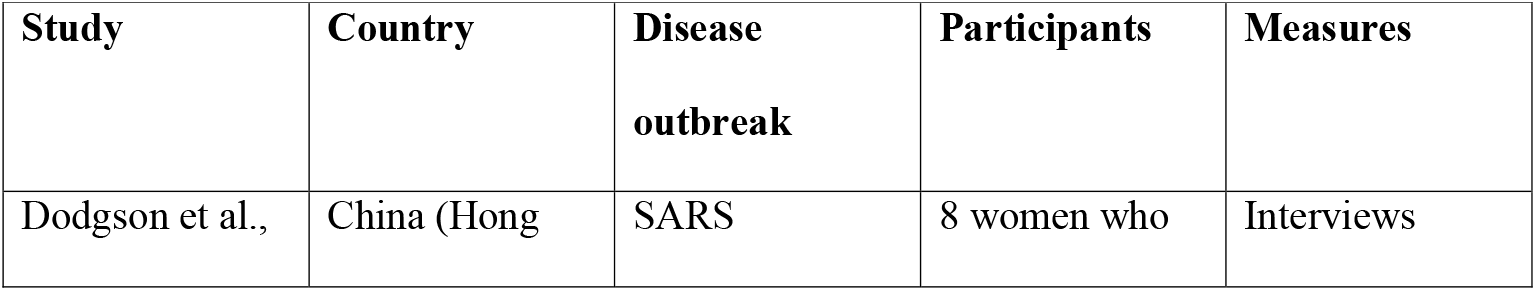
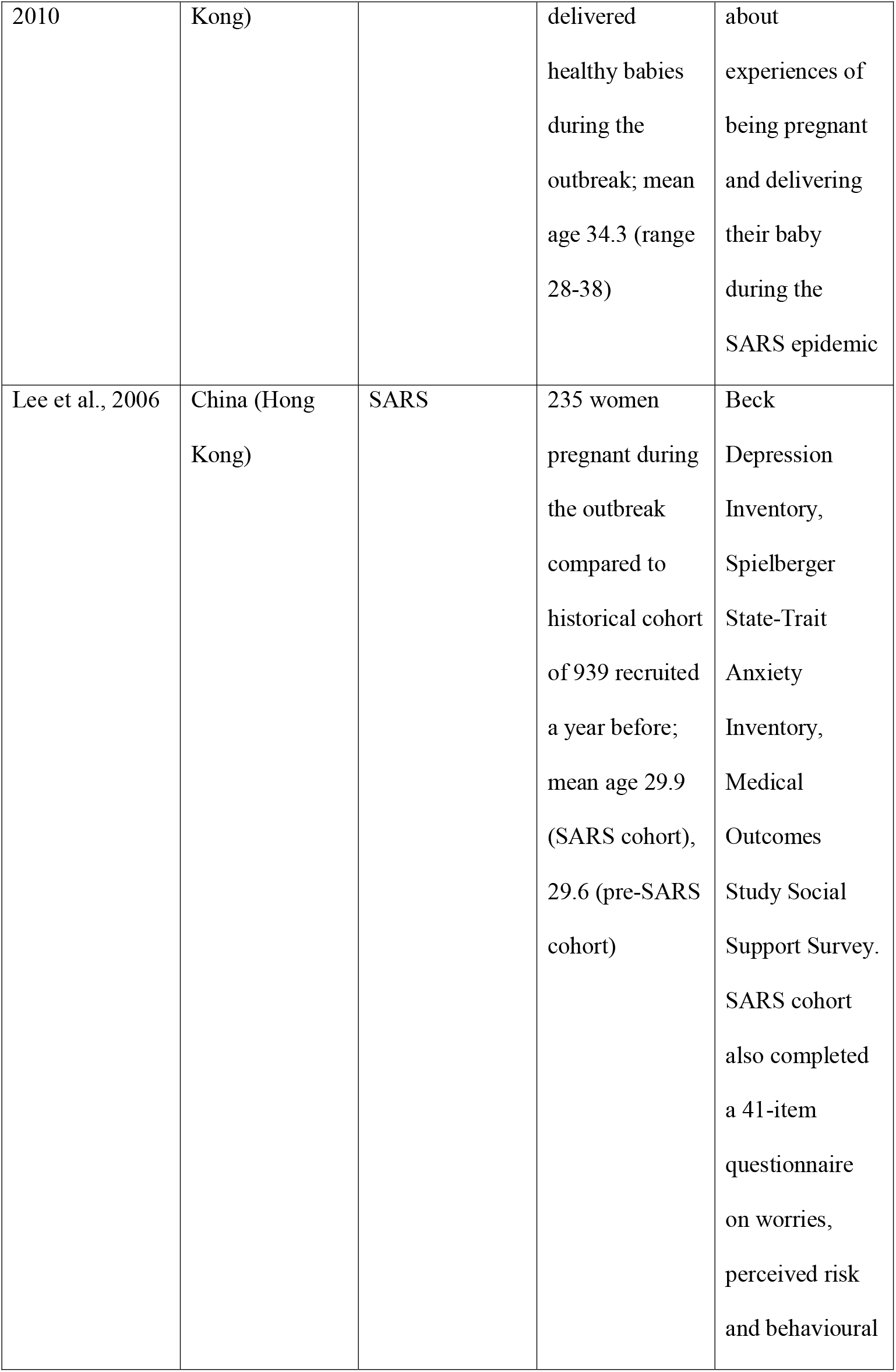
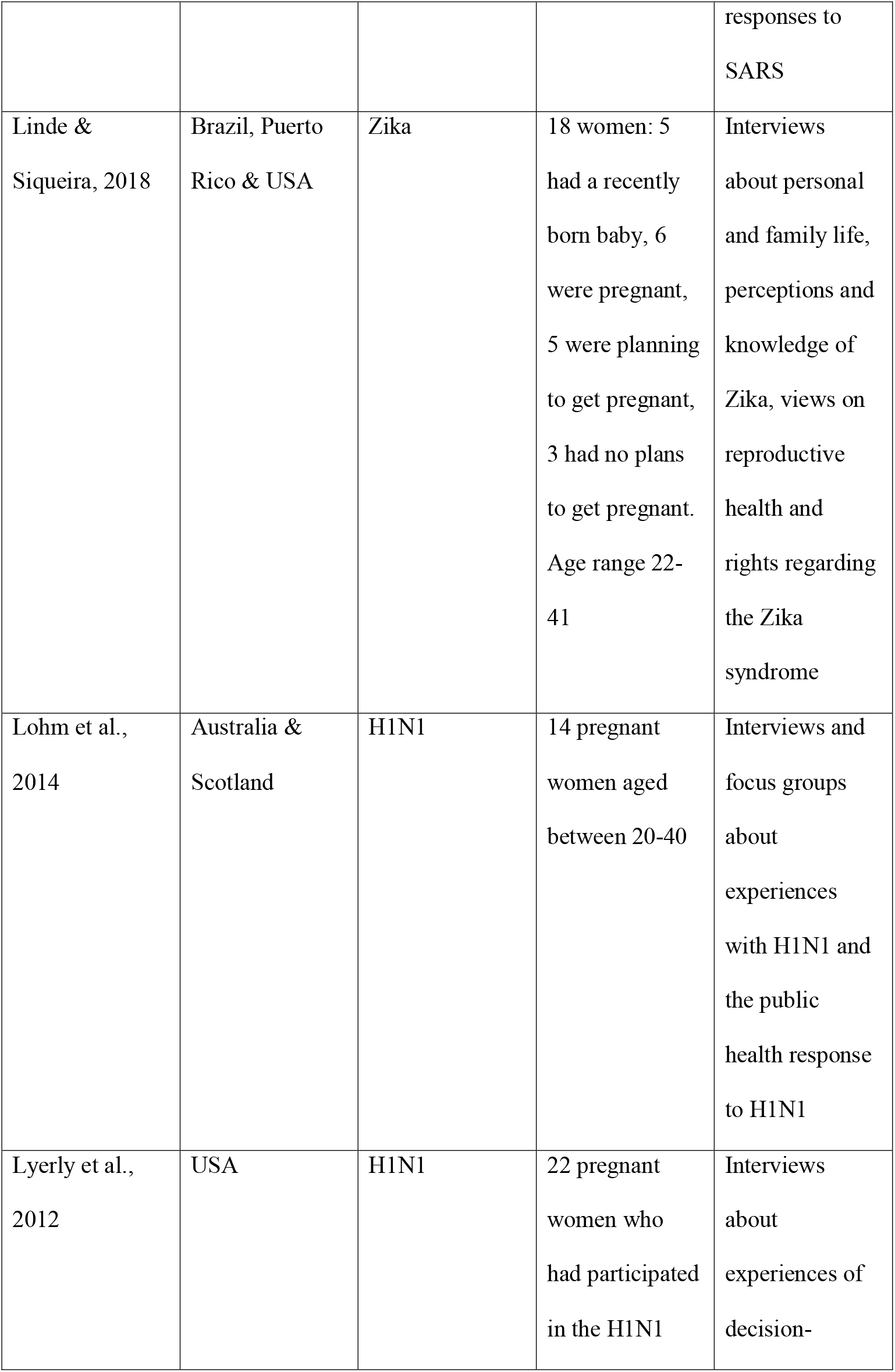
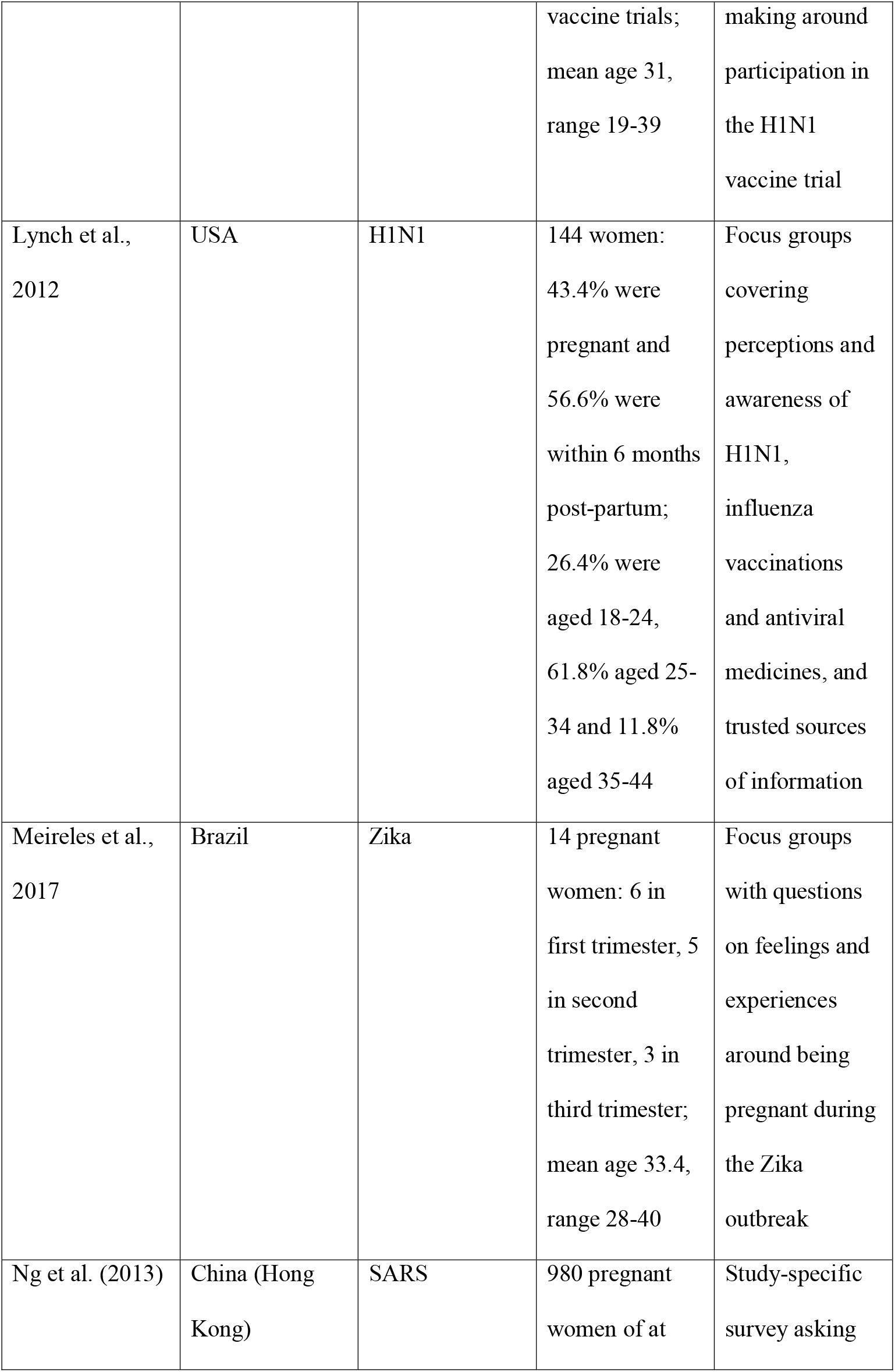
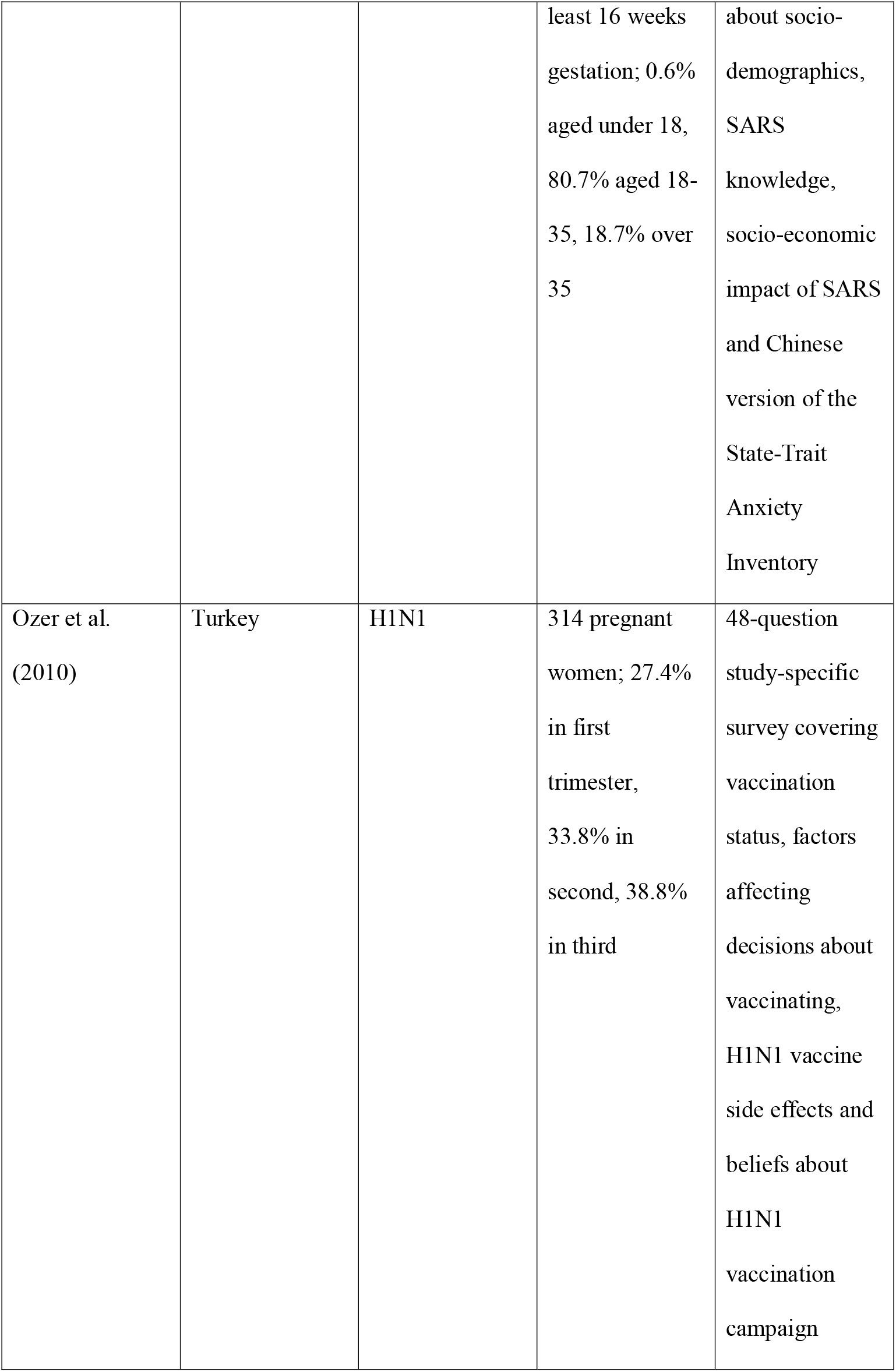
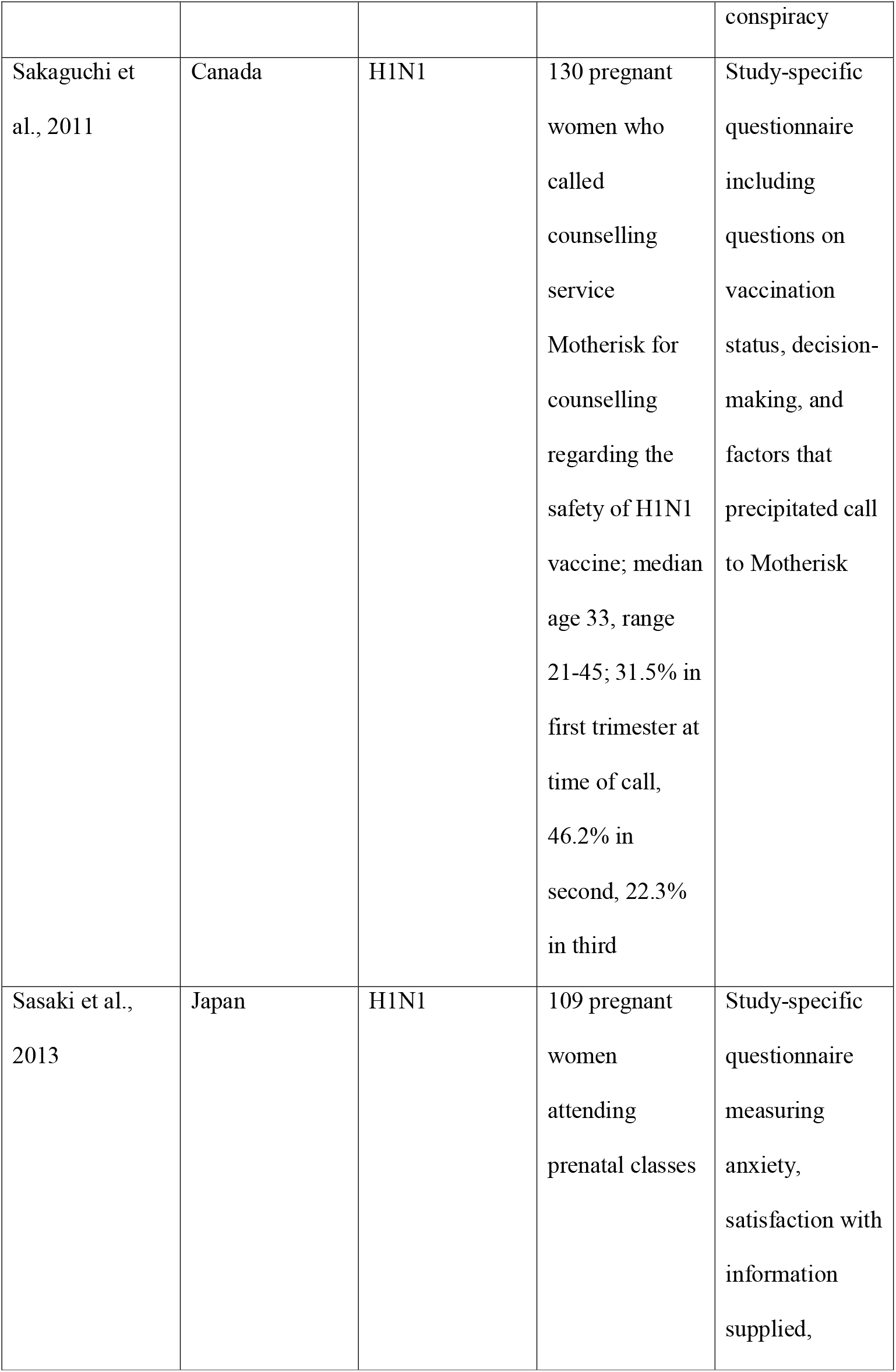
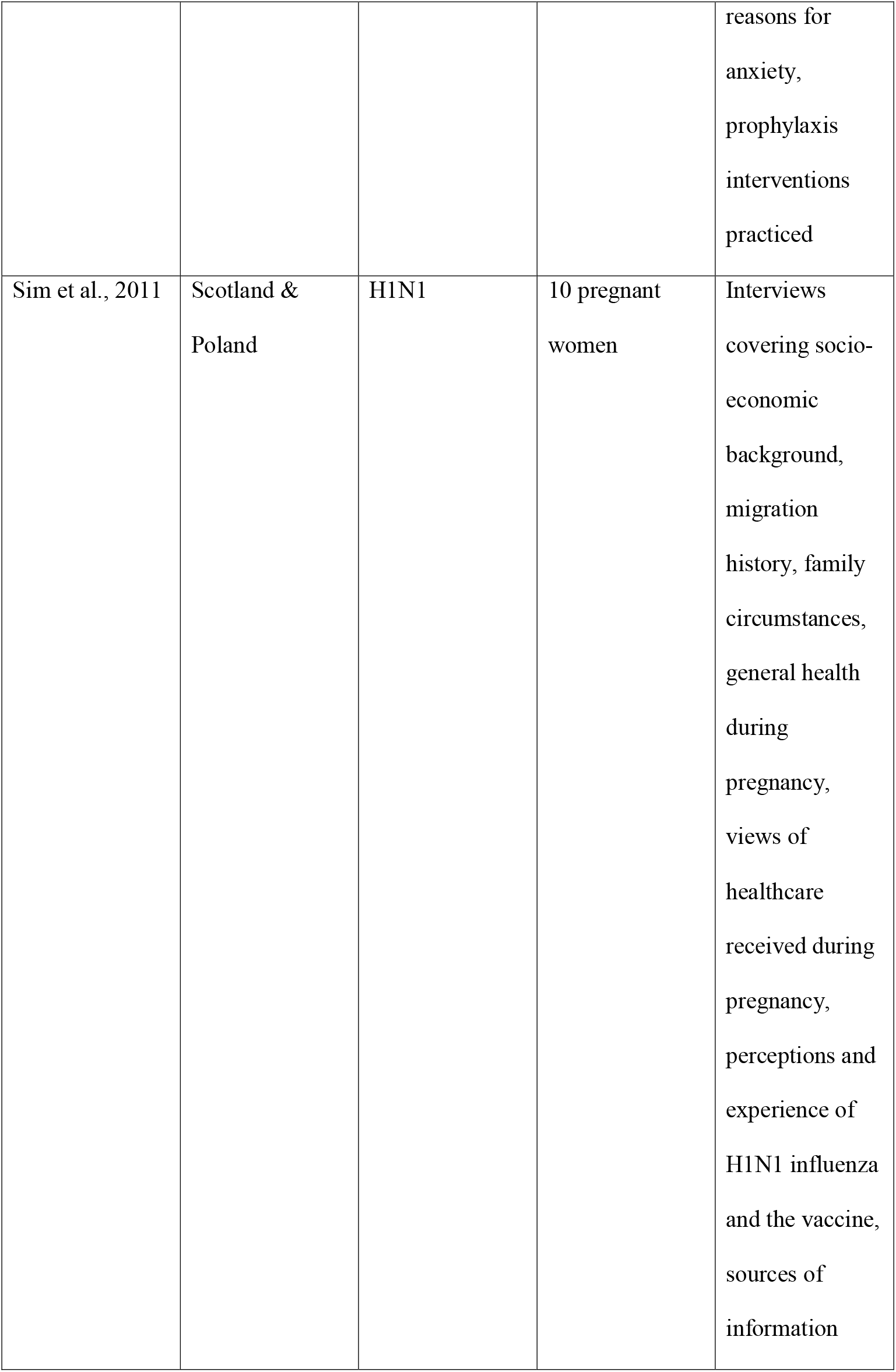
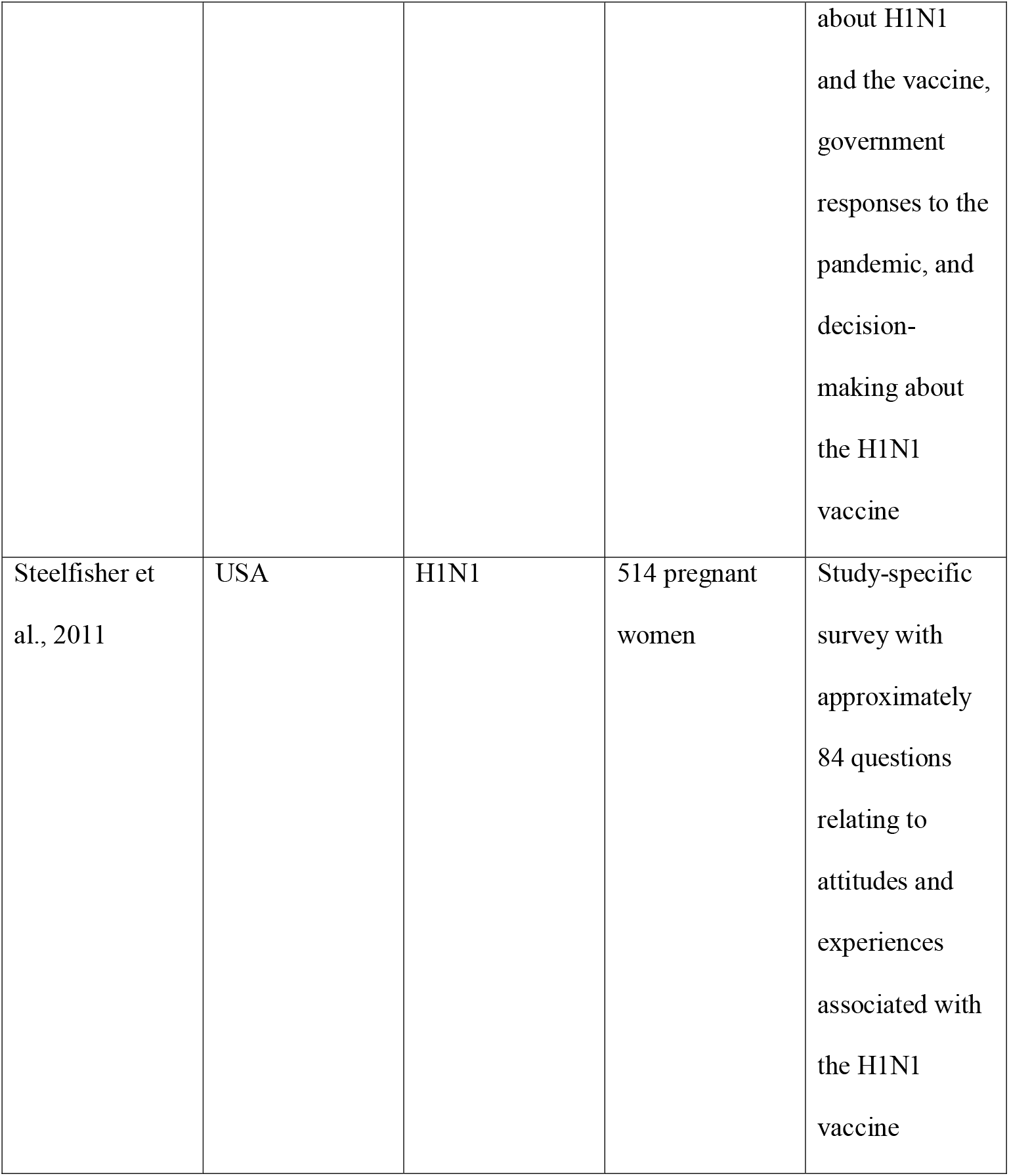
Study characteristics of included articles

**Table II.**
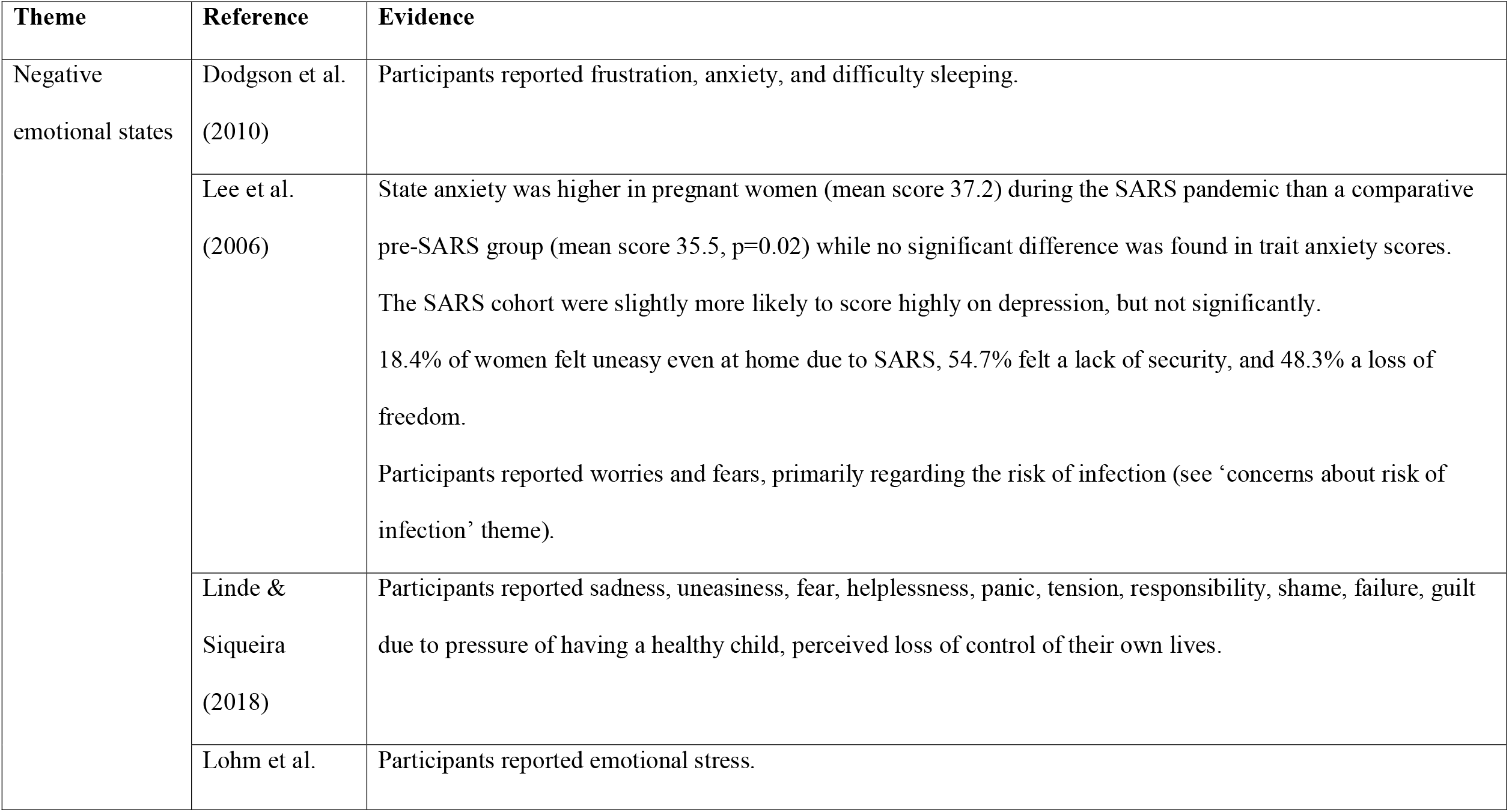
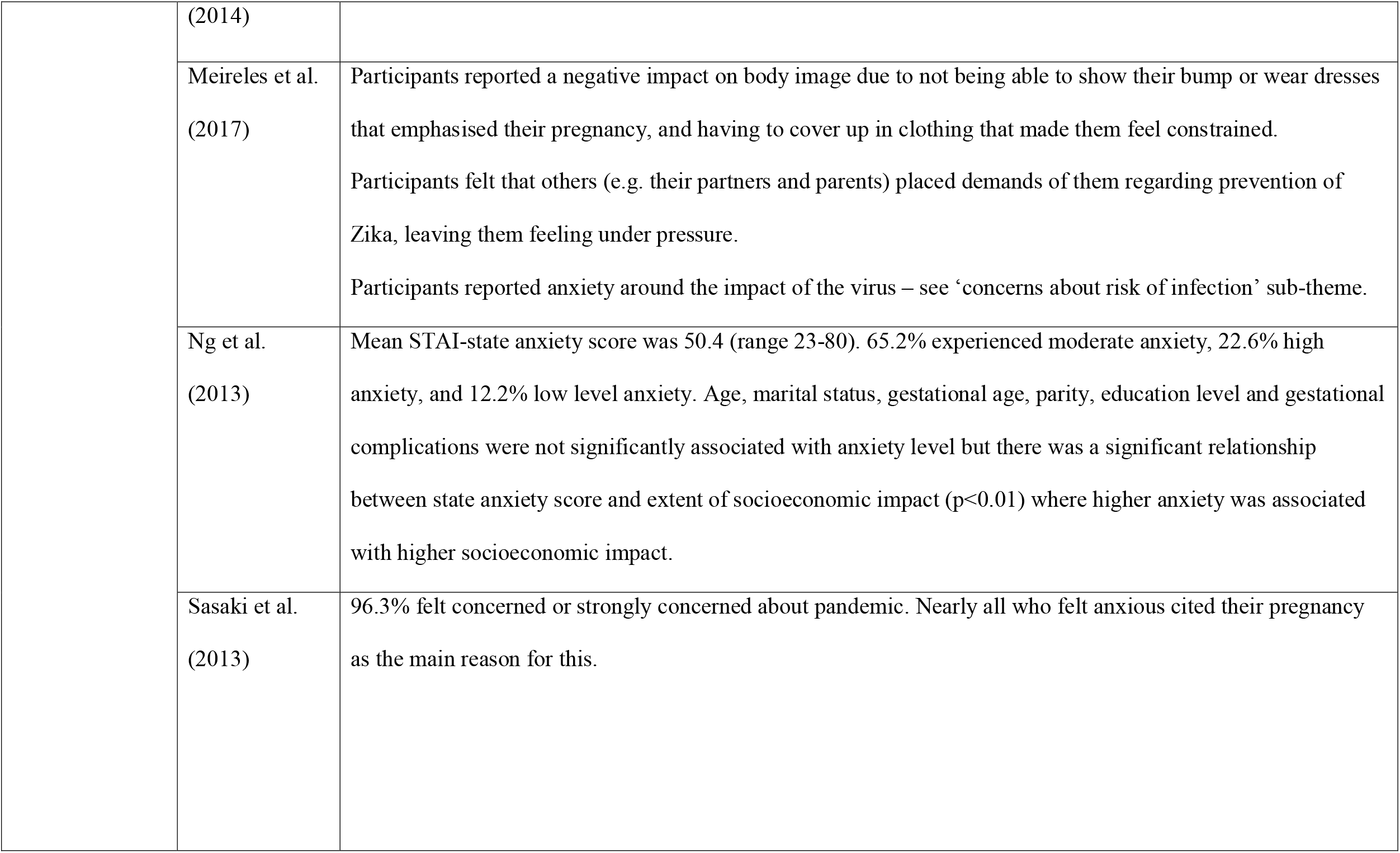
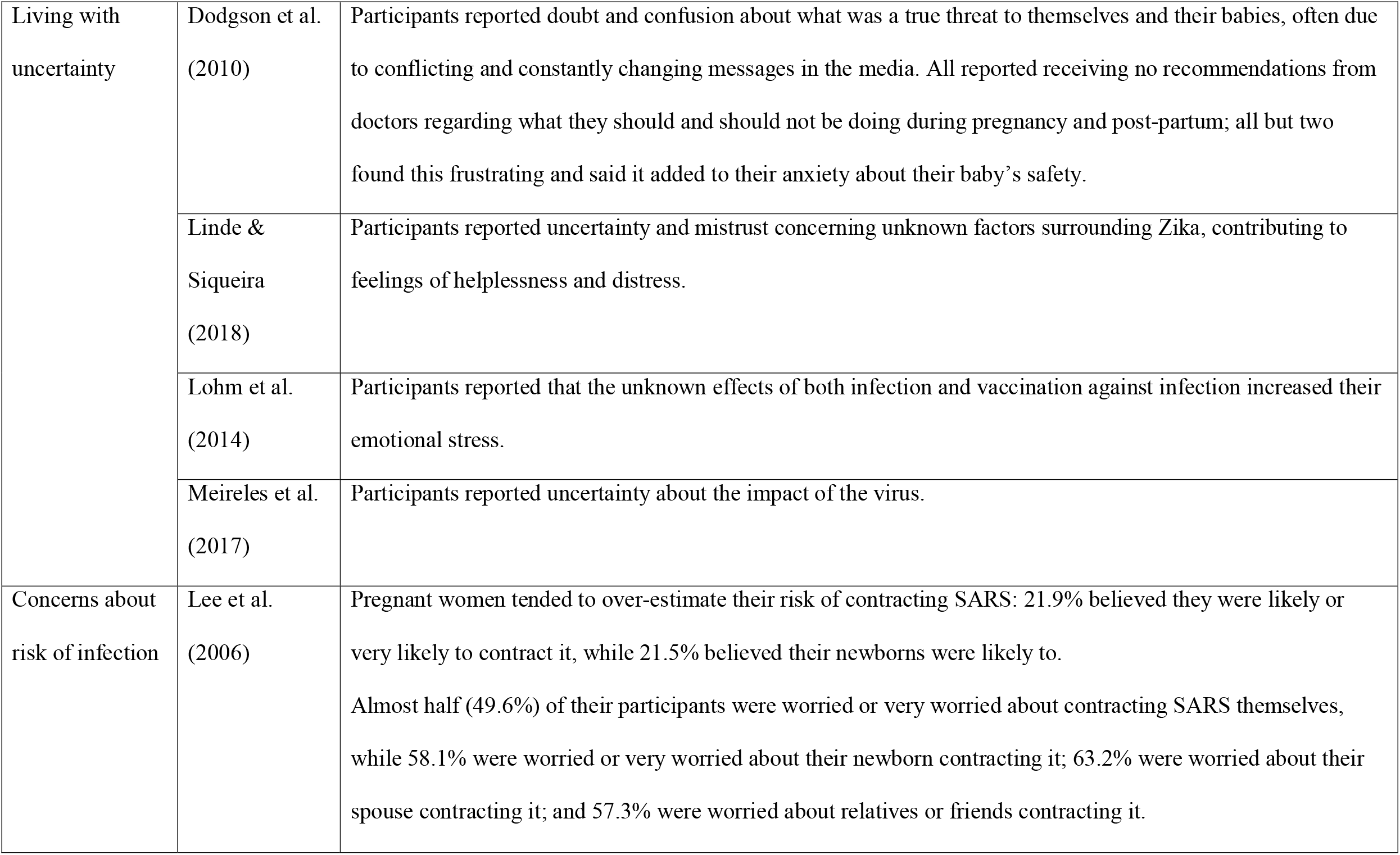
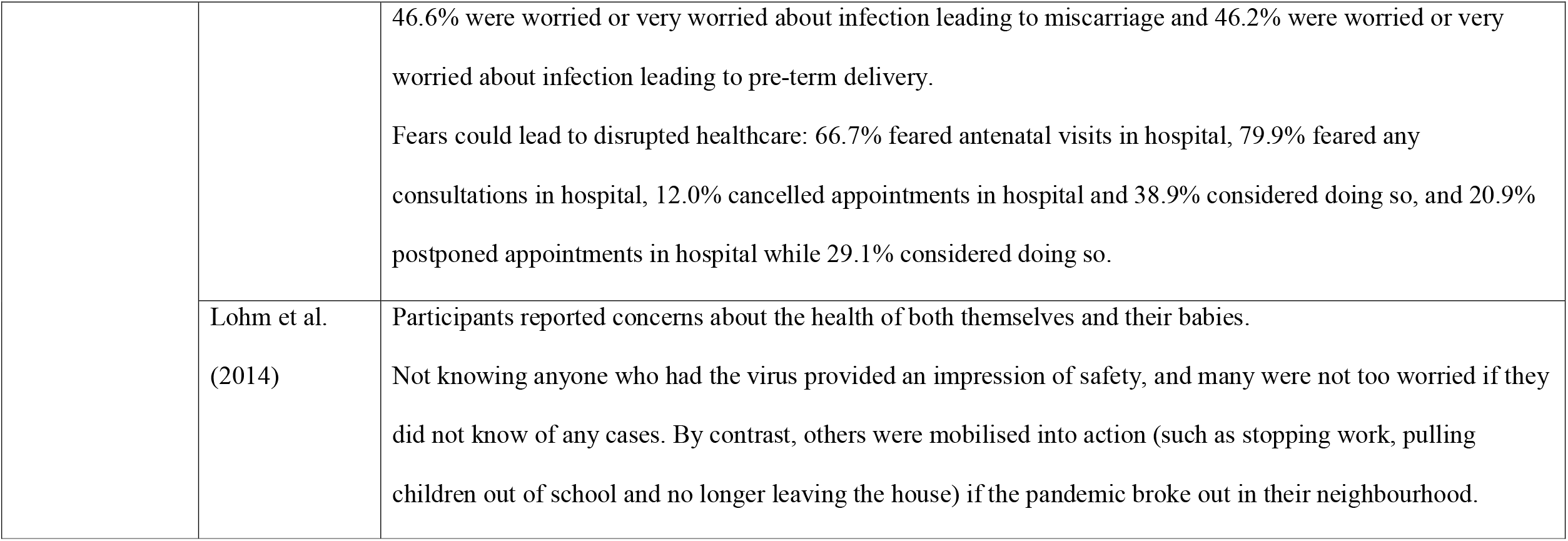
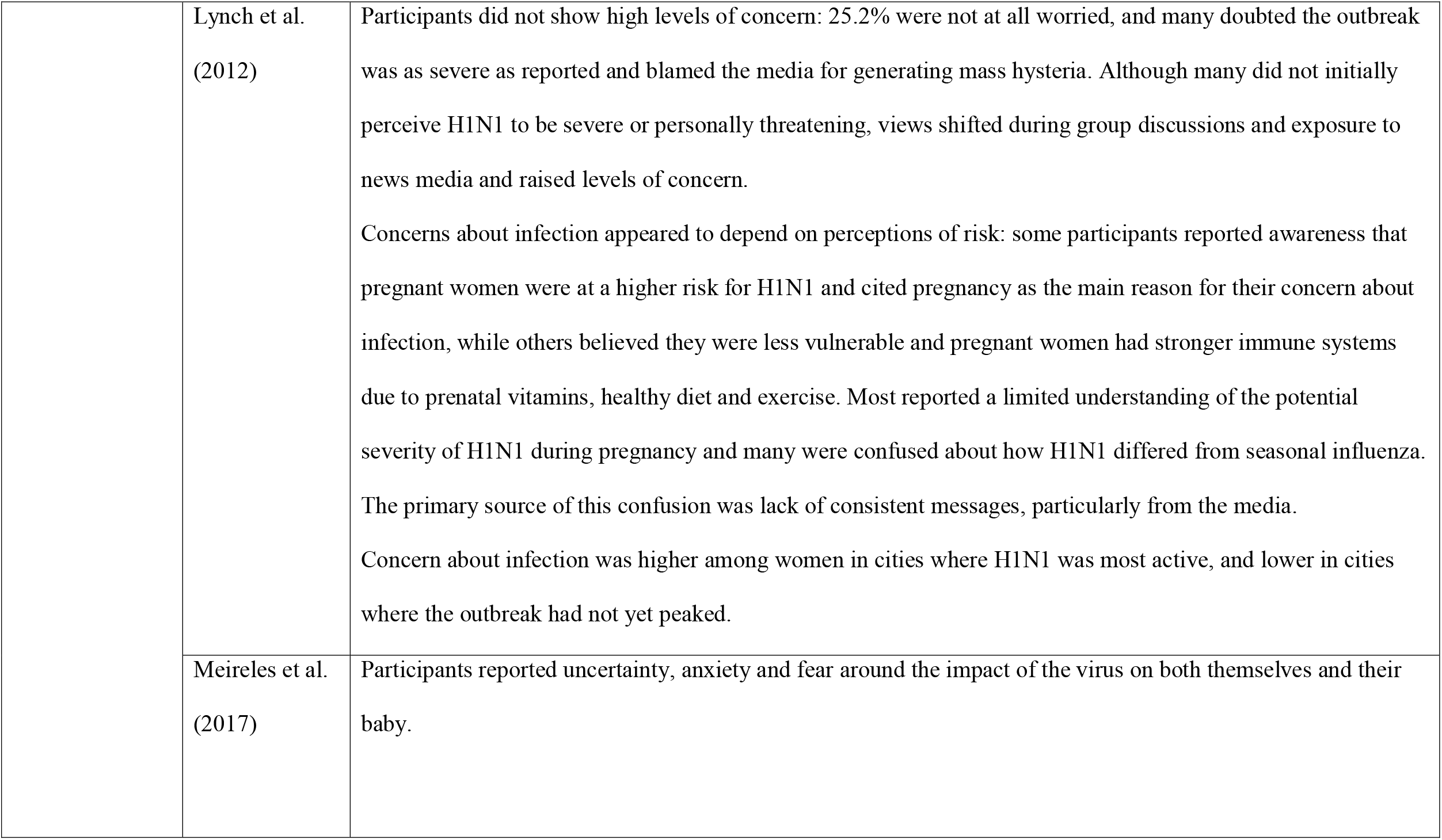
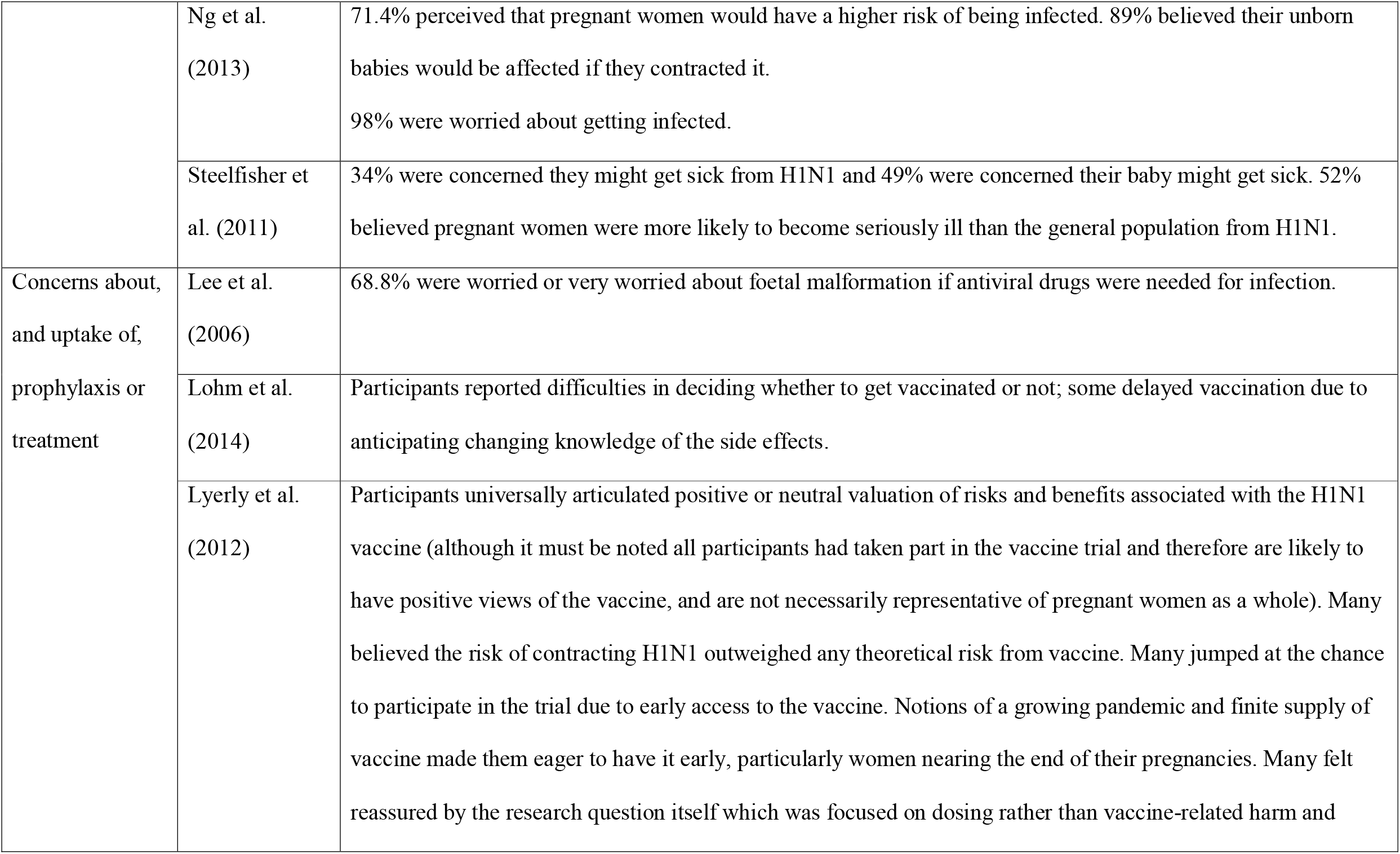
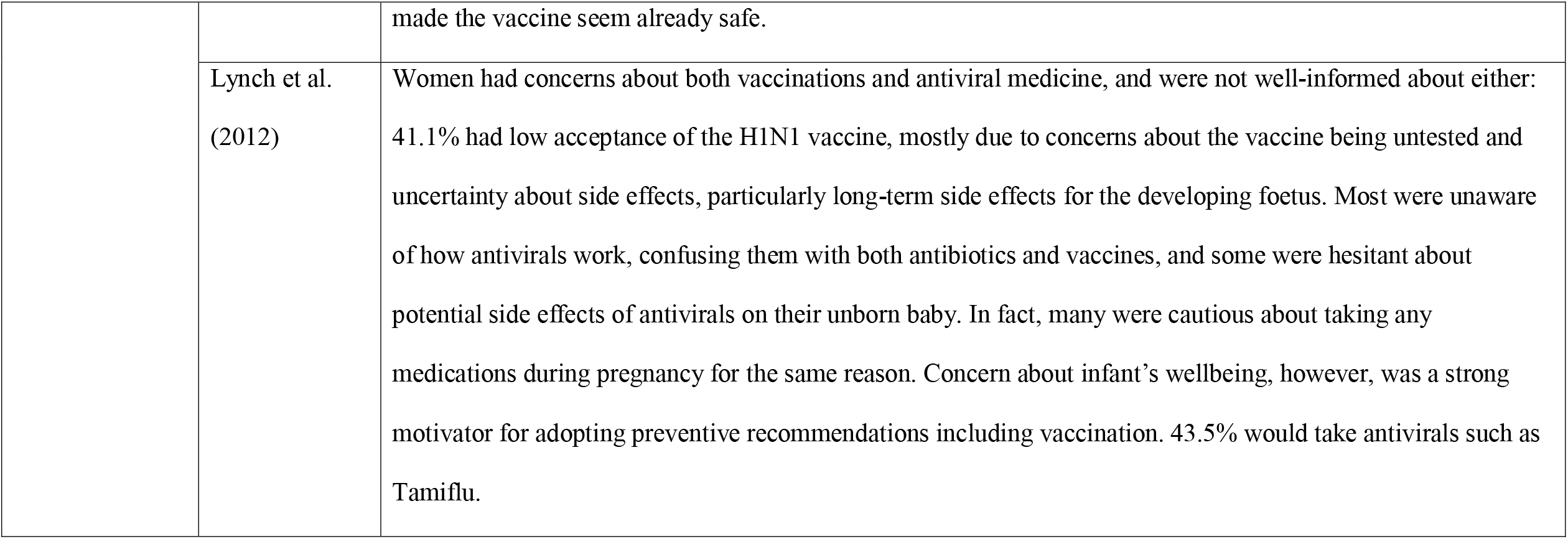
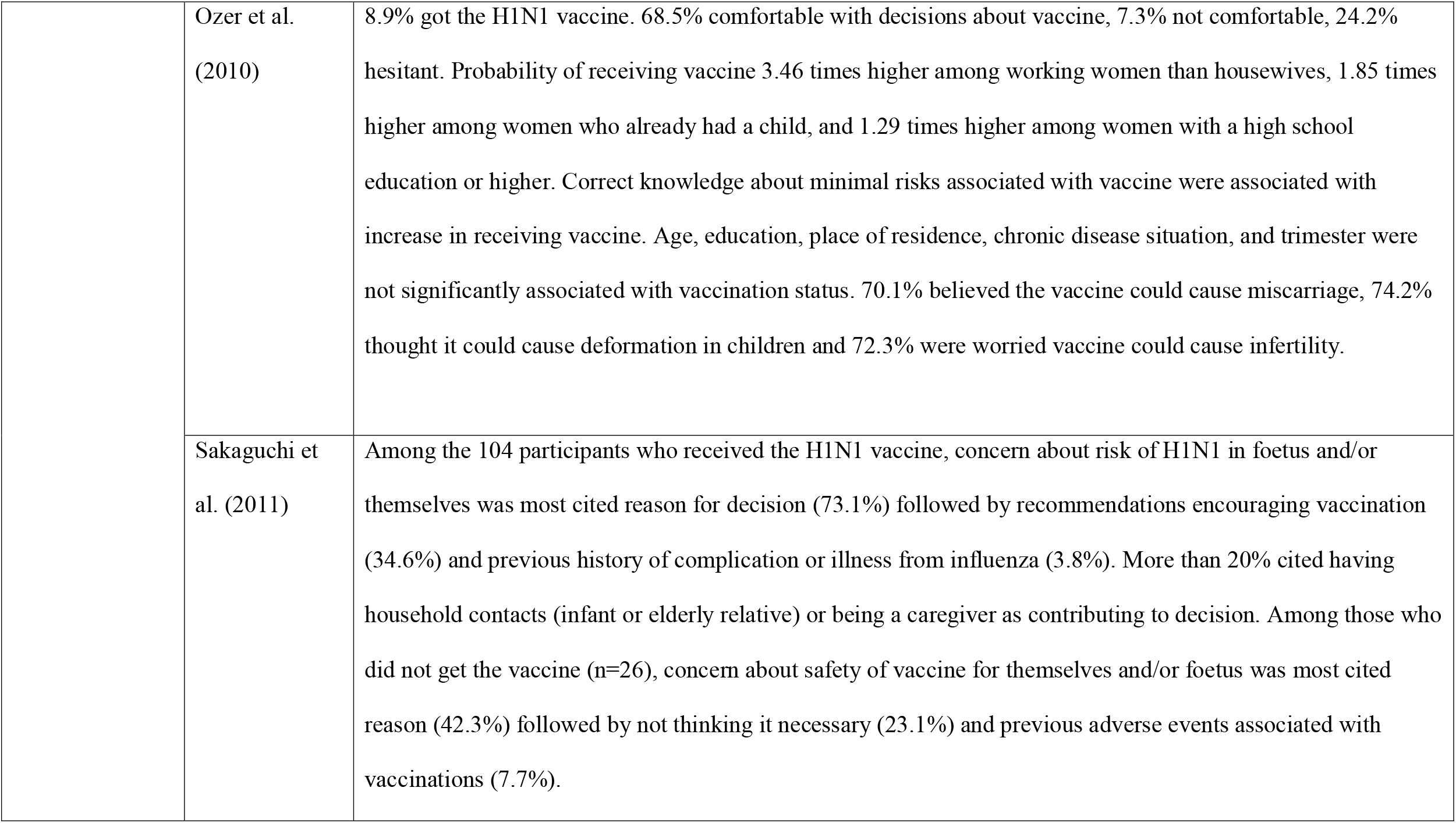
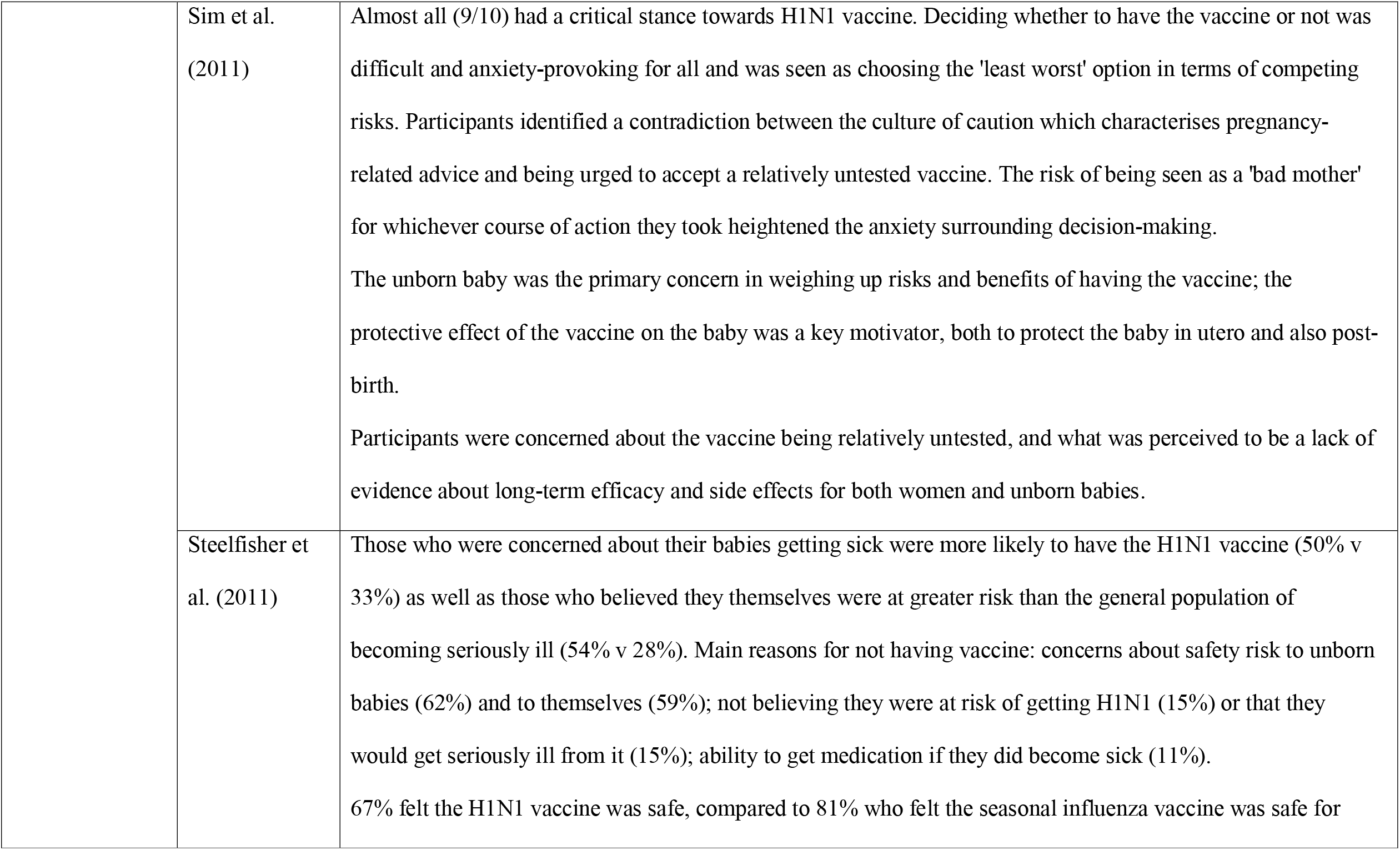
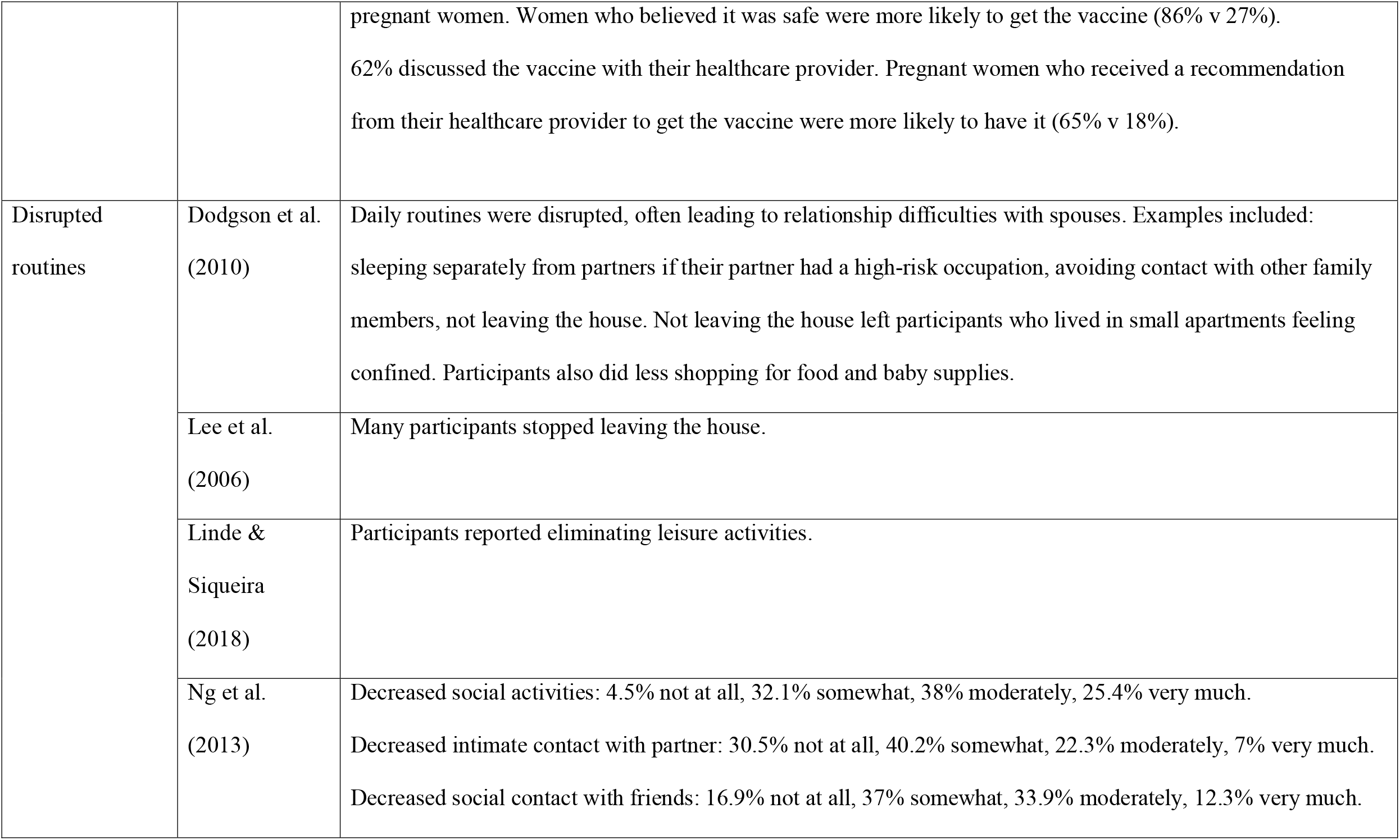
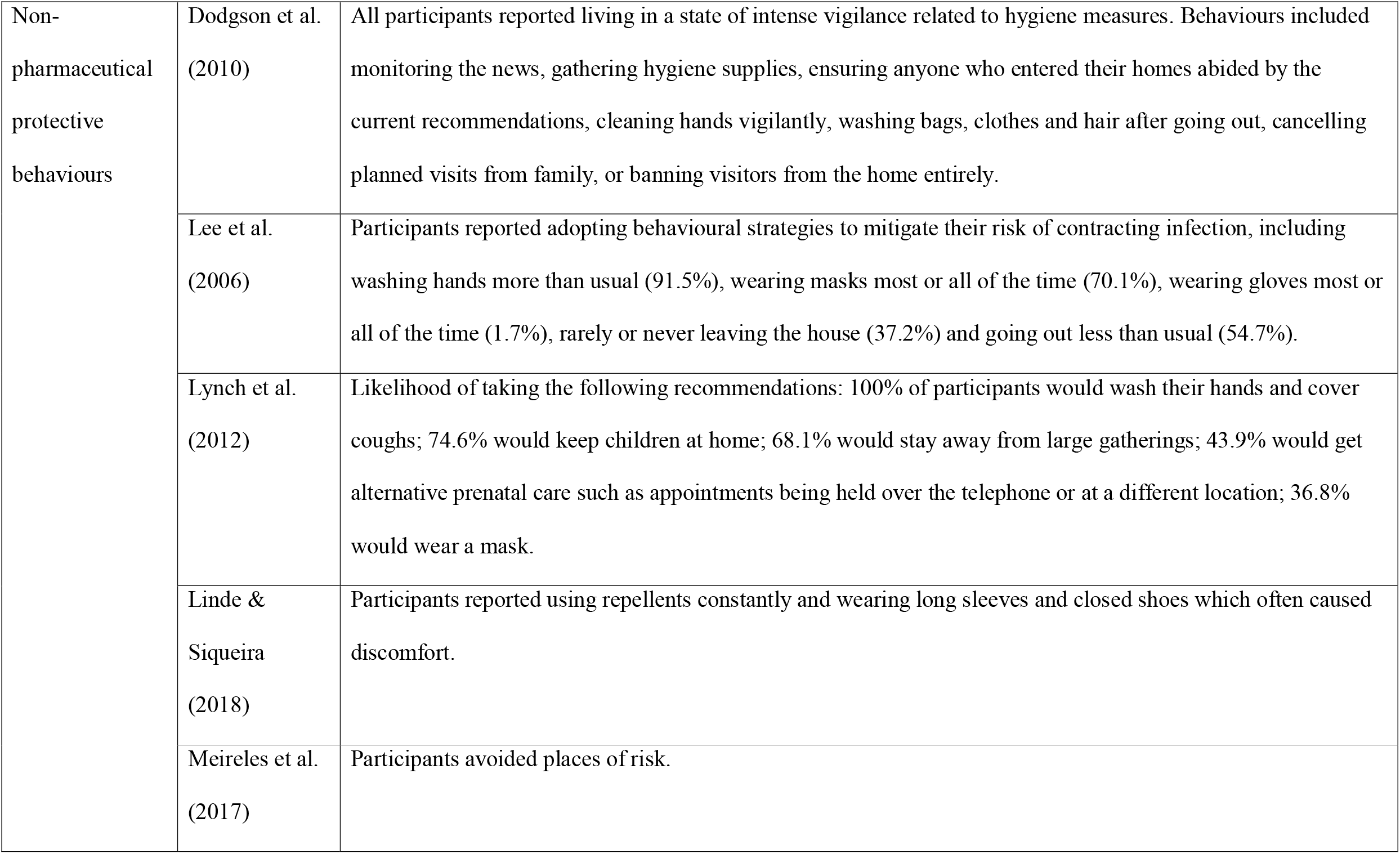
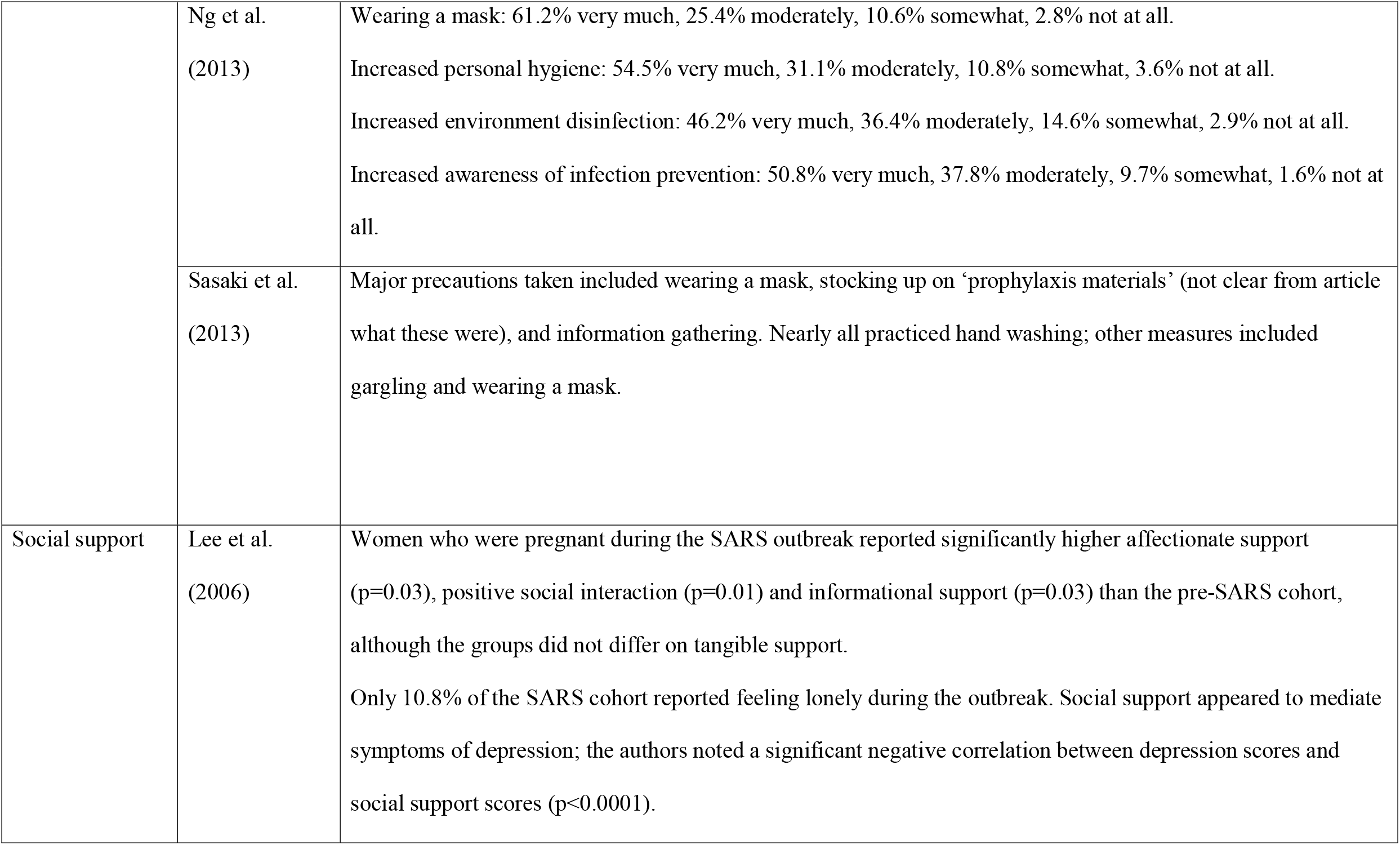
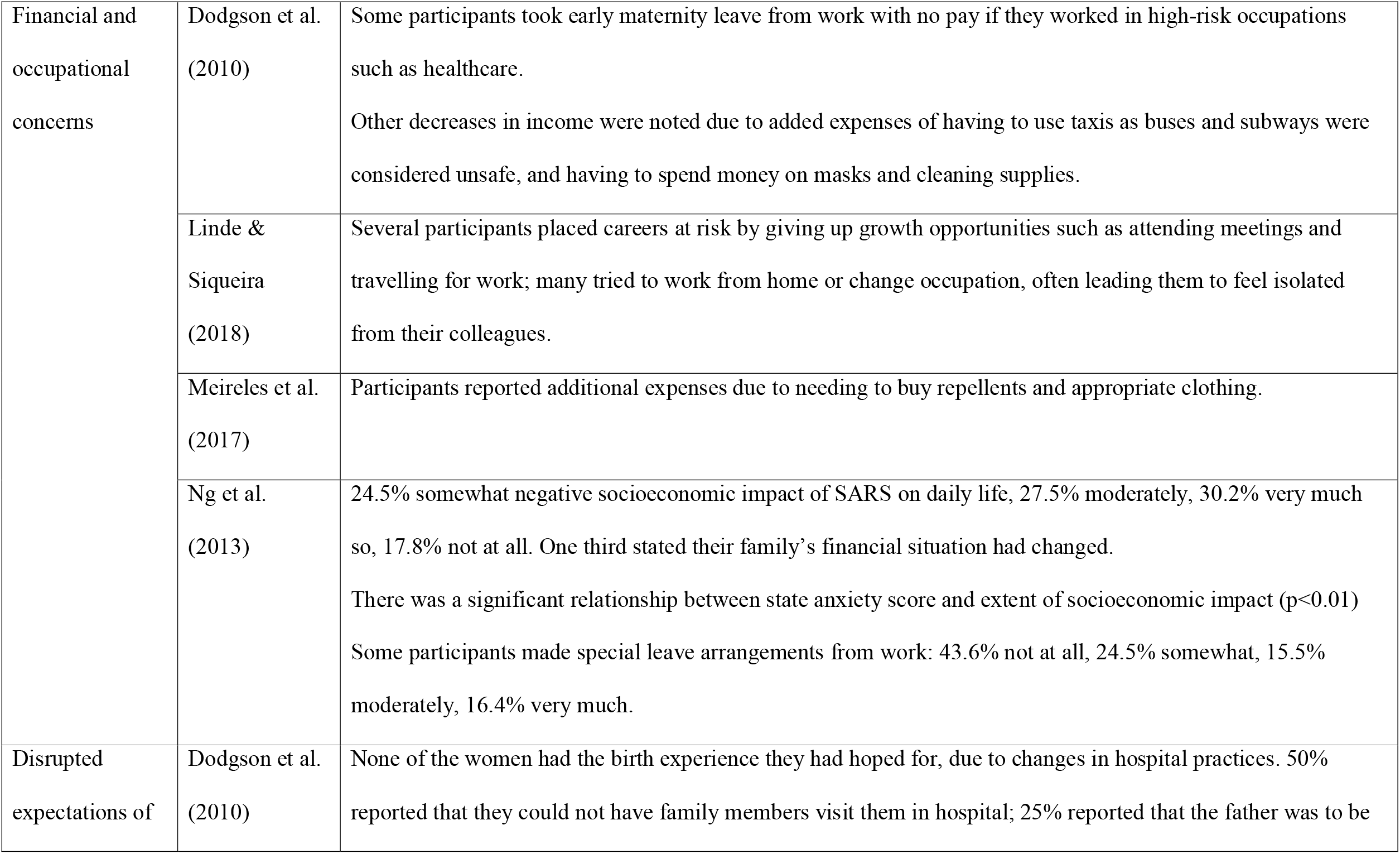
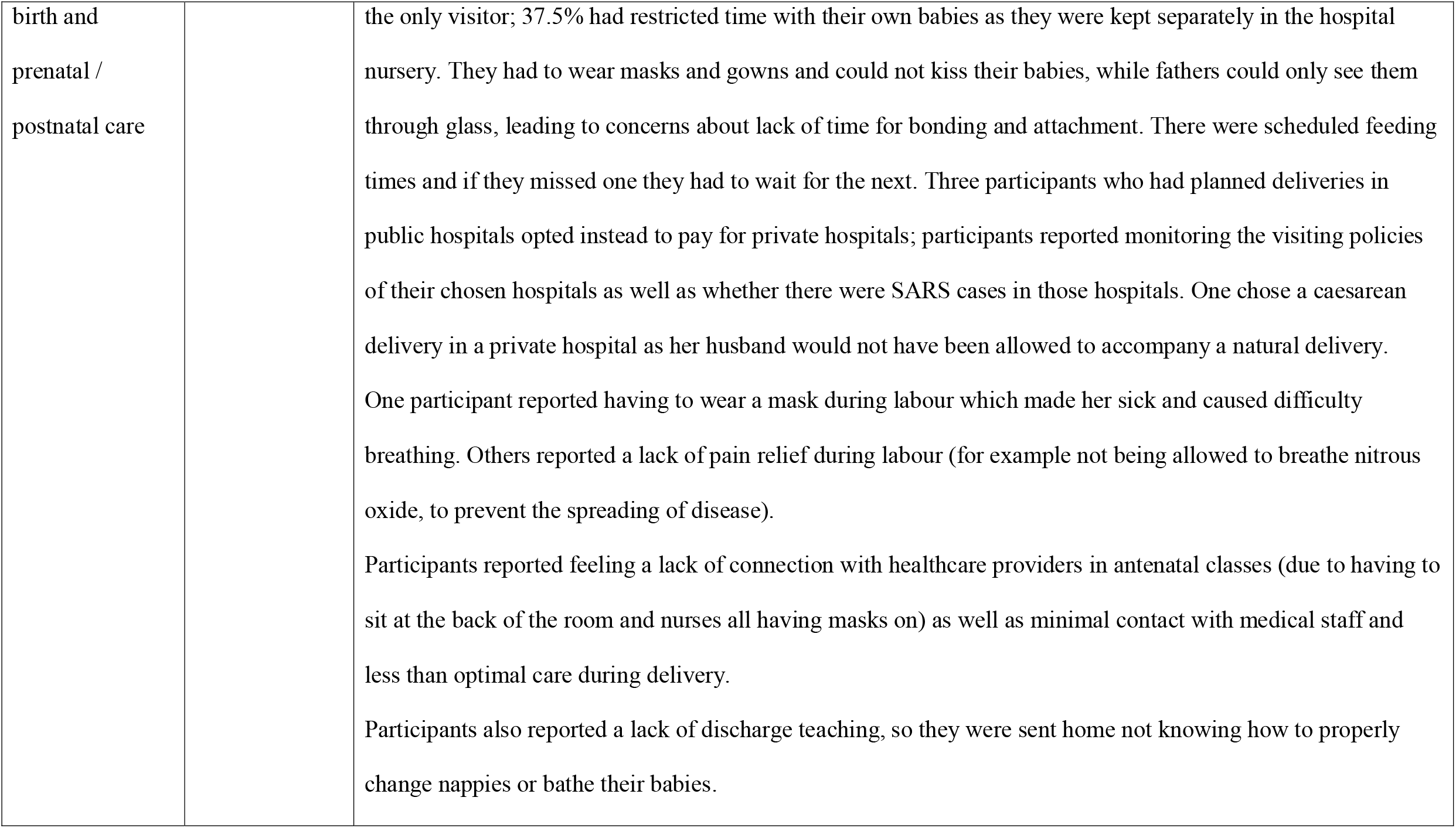
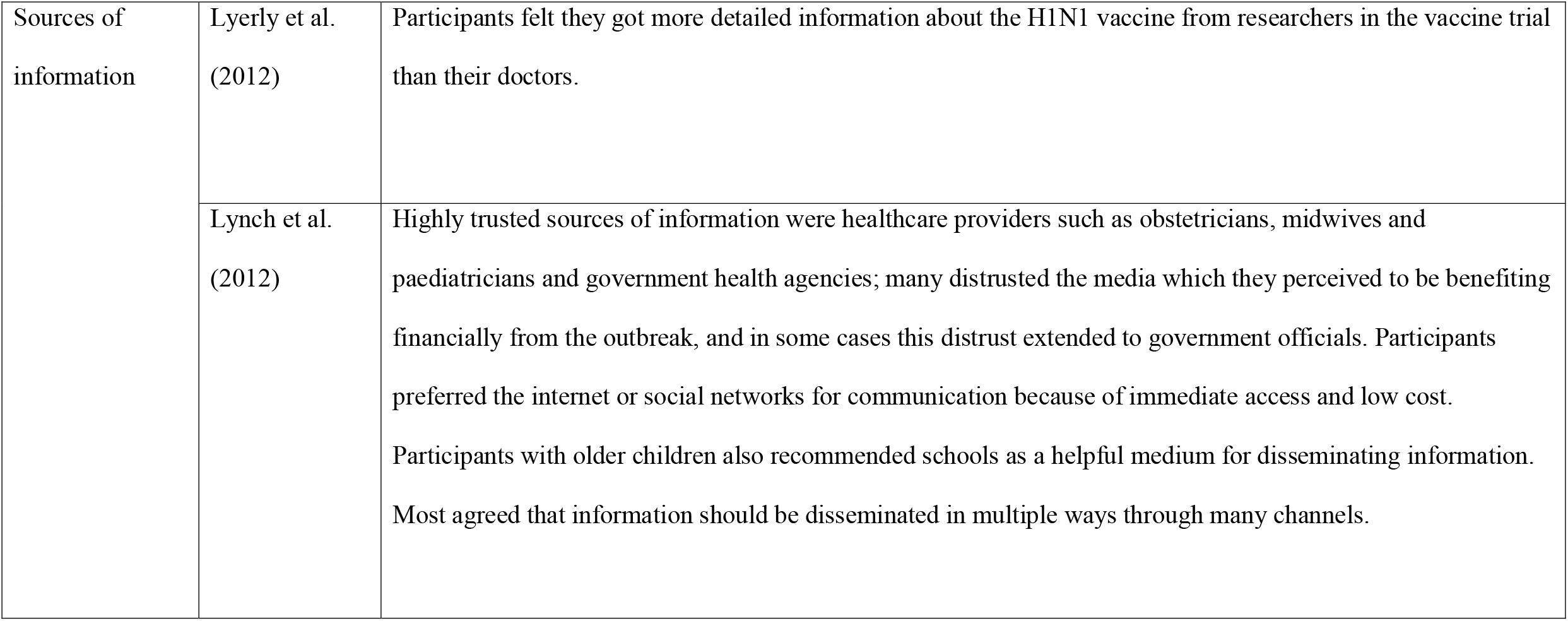
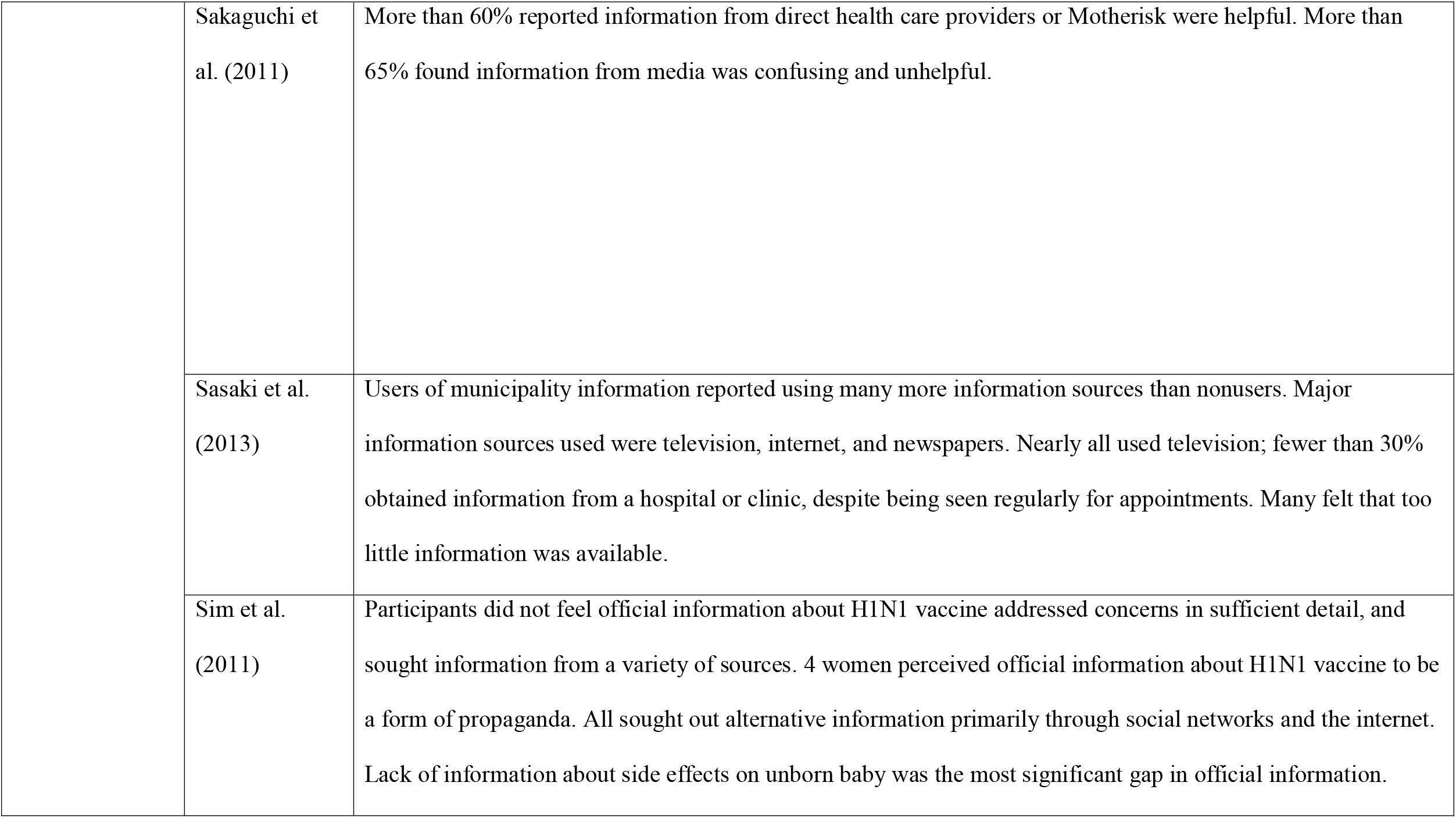
Themes emerging from included studies

### Negative emotional states

Two studies measured state anxiety: Lee et al. (2006) found that state anxiety was significantly higher in pregnant women during the SARS pandemic than a comparative pre-SARS group (p=0.02), and Ng et al. (2013) found that 22.6% of their participants reported high anxiety while 65.2% experienced moderate anxiety. Lee et al. (2006) also found that their SARS cohort were slightly, but not significantly, more likely to score highly on depression. Other negative emotional states included sadness, uneasiness, fear, panic, tension, loss of control of life, shame, failure, and guilt due to the pressure of having a healthy child (Linde & Siqueira, 2018); unease even when at home, feeling a lack of security and a loss of freedom (Lee et al., 2006); stress (Lohm et al., 2014); frustration, anxiety and sleep problems (Dodgson et al., 2010); concern and anxiety about the pandemic situation (Sasaki et al., 2013); pressure from others regarding prevention of infection (Meireles et al., 2017); and negative body image due to having to cover up in protective clothing that made them feel constrained (Meireles et al., 2017).

### Living with uncertainty

Participants in four studies reported living with uncertainty, mostly due to doubt and confusion about the risk to their health and that of their baby (Dodgson et al., 2010; Linde & Siqueira, 2018; Lohm et al., 2014; Meireles et al., 2017). Uncertainty was worsened by conflicting and rapidly changing media messages and not receiving recommendations from doctors regarding what mothers should and should not be doing during pregnancy and post-partum (Dodgson et al., 2010).

### Concerns about risk of infection

Six studies reported that participants expressed concerns about the health of both themselves and their babies (Lee et al., 2006; Lohm et al., 2014; Lynch et al., 2012; Meireles et al., 2017; Ng et al., 2013; Steelfisher et al., 2011), including beliefs that infection could lead to miscarriage or pre-term delivery (Lee et al., 2006) and fears that infection could affect healthcare due to causing participants to cancel antenatal appointments to avoid visiting the hospital (Lee et al., 2006). Levels of concern differed between studies: a study from Hong Kong (Lee et al., 2006) found that approximately half of their participants were worried or very worried about contracting SARS themselves, while in a study from China, Ng et al. (2013) found that 98% of participants were worried about getting infected; Lynch et al. (2012) found that their participants in the USA did not show high levels of concern and over a quarter were not at all worried, doubting the outbreak was as severe as reported and blaming the media for generating hysteria.

Our data suggests that concerns about infection may depend on perceptions of risk: Lee et al. (2006) and Ng et al. (2013) found that pregnant women may have a tendency to over-estimate their risk of contracting infectious diseases: Lee et al. (2006) found that 21.9% of participants believed they were likely or very likely to contract SARS, while 21.5% believed their newborns were likely to. In Ng et al.’s (2013) study, 71.4% perceived that pregnant women had a higher risk of being infected and 89% believed their unborn babies would be affected if they contracted it. Conversely, while a minority of Lynch et al.’s (2012) participants reported awareness that pregnant women were at a higher risk for H1N1 and cited pregnancy as the main reason for their concern about infection, many believed they were less vulnerable as they believed pregnant women had stronger immune systems due to prenatal vitamins, healthy diet and exercise. Most participants in this study reported a limited understanding of the severity of H1N1 and were confused about how it differed from seasonal influenza. It should be noted that though many participants in this study did not initially perceive H1N1 to be severe or personally threatening, their views shifted during focus group discussions and exposure to news media, and increased their concerns.

Perception of risk was affected by proximity to the disease outbreak: lower perceived risk was observed in participants who did not know anyone with the virus (Lohm et al., 2014) or who lived in cities where the outbreak had not yet peaked (Lynch et al., 2012). Conversely, higher perceived risk was observed in participants who lived in neighbourhoods where the infection was higher (Lohm et al., 2014; Lynch et al., 2012).

### Concerns about, and uptake of, prophylaxis or treatment

Seven studies reported that some pregnant women expressed concerns about antiviral drugs (Lee et al., 2006; Lynch et al., 2012) or vaccinations (Lohm et al., 2014; Lynch et al., 2012; Ozer et al., 2010; Sakaguchi et al., 2011; Sim et al., 2011; Steelfisher et al., 2011). Concerns about antivirals were mostly due to being unaware of how they work and being hesitant about potential side effects on unborn babies (Lynch et al., 2012) while concerns about vaccines were mostly due to concerns about the vaccine being untested and uncertainty about side effects, particularly long-term side effects for the developing foetus (Lynch et al., 2012; Ozer et al., 2010; Sakaguchi et al., 2011; Sim et al., 2011; Steelfisher et al., 2011). Reasons for lack of uptake of vaccines included anticipating changing knowledge of the side effects (Lohm et al., 2014), not thinking it necessary (Sakaguchi et al., 2011), and previous adverse vaccination effects (Sakaguchi et al., 2011). Worryingly, over 70% of Ozer et al.’s (2010) participants believed the H1N1 vaccine could cause miscarriage, deformation in children or infertility. Many of Lynch et al.’s (2012) participants reported being cautious about taking any medications at all during pregnancy in case of side effects; similarly, Sim et al.’s (2011) participants noted the contradiction between the culture of caution characterising most pregnancy-related health advice and being urged to have a relatively untested vaccine. Furthermore, some women were concerned about being seen as a bad mother no matter which decision they made about having the vaccine.

However, participants also reported various motivators for receiving vaccines: the primary motivator appeared to be concerns about infants’ wellbeing (Lynch et al., 2012; Sakaguchi et al., 2011; Sim et al., 2011) along with recommendations encouraging vaccination, previous history of complication or illness from influenza (3.8%), and having contact with vulnerable people such as infants or elderly relatives or being a caregiver (Sakaguchi et al., 2011). Factually correct knowledge about the minimal risks of vaccine were associated with increased likelihood of receiving the vaccine (Ozer et al., 2010). One study from the USA (Lyerly et al., 2012) reported entirely positive or neutral valuations of the H1N1 vaccine, with no negative perceptions voiced by participants, although it must be noted all participants had chosen to receive the vaccine as part of the vaccine trial and are not necessarily representative of pregnant women as a whole. Participants in this study believed the risk of contracting H1N1 outweighed any theoretical risk from the vaccine.

### Disrupted routines

Pregnant women’s daily routines, social lives and leisure activities were disrupted as they tried to eliminate the risk of contracting the diseases (Dodgson et al., 2010; Lee et al., 2006; Linde & Siqueira, 2018; Ng et al., 2013). Some did not leave their homes at all (Dodgson et al., 2010; Lee et al., 2006), which could lead to them feeling confined especially when living in a small apartment (Dodgson et al., 2010). Relationships with spouses could be affected, for example due to decreased intimate contact (Ng et al., 2013) and sleeping separately due to fear of infection (Dodgson et al., 2010).

### Non-pharmaceutical protective behaviours

Participants in several studies reported living in a state of vigilance related to hygiene measures and adopting new behaviours to mitigate their risk of contracting infection. Behaviours included monitoring the news and information-gathering (Dodgson et al., 2010; Sasaki et al., 2013); avoiding places of risk (Dodgson et al., 2010; Lynch et al., 2012; Meireles et al., 2012); gathering hygiene supplies (Dodgson et al., 2010); cleaning hands vigilantly (Dodgson et al., 2010; Lee et al., 2006; Lynch et al., 2012; Sasaki et al., 2013); washing bags, clothes and hair after leaving the house (Dodgson et al., 2010); wearing masks (Lee et al., 2006; Lynch et al., 2012; Ng et al., 2013; Sasaki et al., 2013); stocking up on prophylaxis materials (Sasaki et al., 2013); and cancelling planned visits from family or banning visitors to the home altogether (Dodgson et al., 2010). With regards to the Zika virus, participants reported using insect repellents constantly and wearing long sleeves and closed shoes which often caused discomfort (Linde & Siqueira, 2018).

### Social support

Lee et al. (2006) found that women pregnant during the SARS outbreak reported significantly higher affectionate support (p=0.03), positive social interaction (p=0.01) and informational support (p=0.03) than the pre-SARS cohort, although the groups did not differ on tangible support. In this study, only 10.8% of the SARS cohort reported feeling lonely during the outbreak; there is no comparable figure for the pre-SARS cohort as the question on loneliness was only asked to the SARS cohort. Social support appeared to mediate symptoms of depression; the authors noted a significant negative correlation between depression scores and social support scores (p<0.0001).

### Financial and occupational concerns

In Ng et al.’s (2013) study, over a third of participants reported their family’s financial situation had been negatively affected as a result of the outbreak. Participants reported increased expenses due to using taxis since buses and subways were considered unsafe (Dodgson et al., 2010) and having to buy supplies to mitigate their risk of infection, such as masks and cleaning supplies (Dodgson et al., 2010) or insect repellents and clothing (Meireles et al., 2017).

Some participants took early maternity leave from work and forfeited pay if they worked in high-risk occupations such as healthcare (Dodgson et al., 2010), or made special leave arrangements (Ng et al., 2013); others put their careers at risk by giving up career-promoting opportunities which involved attending meetings or travelling (Linde & Siqueira, 2018). In the latter study, many tried to work from home or change occupation, leading them to feel isolated from colleagues.

### Disrupted expectations of birth and prenatal / postnatal care

One study (Dodgson et al., 2010) reported on disrupted expectations of birth, prenatal care and postnatal care. In this study, none of the participants had the birth experience they had hoped for, due to changes in hospital practices such as not allowing family members to visit, not allowing fathers to be present for the birth, having to wear masks during labour, not being allowed pain relief during labour, minimal contact with healthcare staff, only allowing mothers to feed their babies at set times, or restricting new mothers’ time with their babies who were kept separately in the hospital nursery. Mothers had to wear masks and gowns and could not kiss their babies, while fathers could only see them through glass, leading to concerns about lack of time for bonding and attachment. In some cases, these changes in hospital policies led women to change their birth preferences, for example choosing caesarean delivery or delivery in a private hospital.

Prenatal care was also affected: participants reported feeling a lack of connection with healthcare providers in antenatal classes due to having to keep their distance from nurses, and nurses all having masks on. In terms of postnatal care, participants reported a lack of discharge teaching, so they were sent home not knowing how to properly change nappies or bathe their babies.

### Sources of information

Lynch et al. (2012) reported on the preferred sources from which participants got information on the outbreak: healthcare providers such as obstetricians, midwives and paediatricians were generally highly trusted, as well as government health agencies to a lesser extent. Many expressed distrust of the media, which they perceived to be benefiting financially from the outbreak; in some cases this distrust extended to government officials. Participants preferred the internet or social networks for communication because of immediate access and low cost. Participants who were already parents also suggested their children’s schools were a helpful medium for disseminating information about vaccine and antiviral use, to help them make sound decisions. Most agreed that information should be disseminated in multiple ways through many channels. The majority of participants in Sakaguchi et al.’s (2011) study also reported finding information about the H1N1 vaccine from direct health care providers helpful, as well as the Motherisk counselling service (a hospital-based programme providing evidence-based information regarding the safety or risks associated with treatments and infectious diseases during pregnancy); in the same study, more than 65% of participants found that media information was confusing and unhelpful.

Conversely, Sasaki et al. (2013) found that television, internet and newspapers were the most common sources of information about the H1N1 outbreak, while fewer than 30% of participants obtained their information from a hospital or clinic. Many participants felt that too little information was available. Sim et al.’s (2011) participants did not feel that official information about the H1N1 vaccine addressed their concerns, particularly about potential effects on unborn babies, and sought information from a variety of sources such as social networks and the internet. In this study, several participants perceived that the official information provided by the National Health Service was ‘propaganda’.

It is also noteworthy that Lyerly et al. (2012) found that their participants, who had taken part in the H1N1 vaccine trial, felt they had received more information from the trial’s researchers than they had from their doctors.

## Discussion

The results of this review suggest a considerable potential for disease outbreaks to have a negative emotional impact on pregnant women, creating anxiety, distress and fear. Other themes emerging from the review, and also creating a negative impact, include uncertainty; concerns about infection and related health risks to both the self and unborn babies; concerns about prophylaxis or treatment; disrupted routines; financial and occupational concerns; and disrupted expectations of birth and prenatal and postnatal care. Intense vigilance with regards to non-pharmaceutical protective behaviours (such as wearing masks, hand-washing and avoiding contact with others) was frequently reported. Social support may be a protective factor for poor mental health although during an outbreak this may be difficult to access. Sources of disease information included healthcare providers, government health agencies and the media; participants expressed mixed opinions about the trustworthiness of the various sources although healthcare professionals tended to be viewed as the most trustworthy.

This review suggests that being pregnant during a time of an outbreak increases the vulnerability of pregnant women to the associated stress of the situation. Indeed, the few papers in this review with measures of mental health suggested high levels of anxiety in pregnant women during infectious disease outbreaks. While no papers compared anxiety in pregnant and non-pregnant women during a disease outbreak, and it is likely that outbreaks can cause anxiety for all, one study (Lee et al., 2006) compared a pre-SARS group with a group who were pregnant during the SARS outbreak and found that state anxiety was significantly higher during the outbreak; another (Sasaki et al., 2013) found that nearly all pregnant women cited that being pregnant during an outbreak was their primary reason for feeling anxious. This is concerning as previous research suggests that experiencing prenatal stress can lead to adverse birth outcomes (Pais & Pai, 2018). Early identification of mental health issues in perinatal patients is therefore essential; midwives should be aware of pregnant women’s propensity to experience intense anxiety during outbreaks and they should take account of the potential impact of such symptoms on their physical and mental health. Early identification of problems can allow obstetric providers to partner with mental health specialists in order to establish appropriate comprehensive treatment plans (Maher, 2019). Planning and provision of education of, and mental health services specifically for, pregnant women during infectious disease outbreaks is likely to be useful (Ng et al., 2013).

Feelings of uncertainty were cited by participants as a significant stressor in several studies. This is likely to be the case for many people – not just for pregnant women – as stress has been frequently linked to uncertainty across the population as a whole (Grupe & Nitschke, 2013). Again, this is particularly concerning for pregnant women as previous research suggests that uncertainty can cause fear and distress in pregnancy (Melander, 2002) which, as outlined above, could in turn lead to adverse birth outcomes (Pais & Pai, 2018). Pandemics are characterised by uncertainty for everyone, but public health officials and healthcare providers can take steps to reduce this uncertainty as much as possible by ensuring that information provided to the public is timely, accurate, and consistent with information from other sources. This review provided some evidence that the media is perceived as untrustworthy, and so information directly from healthcare providers and official public health organisations appear preferable. The distrust of media reporting noted in other studies may be prevalent across the population as a whole, not just for pregnant women (Goodall et al., 2012). The current outbreak advice is not to watch much media and to seek information only from trusted sources (Williamson et al., 2020); pregnant women can take action themselves to avoid media especially if it causes them to become anxious.

Many participants in the reviewed studies expressed concerns about becoming infected, and the negative effects this could have on their own health and that of their babies; levels of concern differed across studies, and it is possible that these differences in concern about infection could be cultural. Concerns about infection appeared to be related to perceived rather than actual risk, with some participants over-estimating the risk of infection during pregnancy. This highlights the need for timely dissemination of accurate information, and for clinicians to monitor for over-estimation of infectious risk among pregnant women and clear up misconceptions. Where simple advice and reassurance does not work, there may be benefit in brief psychotherapy using a cognitive-behavioural model in order to reduce anxiety and the associated risk of pregnancy complications (Asghari et al., 2016; Goodman et al., 2014).

Concerns about prophylaxis or treatment were prevalent. This is perhaps unsurprising as pregnant women have historically low vaccination rates for seasonal influenza (Yuen & Tarrant, 2014) and pandemic influenza (Fridman et al., 2011, Schmid et al., 2017). The decision about whether to receive vaccines or medications may well be distressing as pregnancy is already a time when women are faced with cultural expectations of motherhood involving eliminating all risk to the foetus and complying with medical advice and any examples of not abiding by advice lead to women being seen as undisciplined or ignorant (Bessett, 2010). It is essential that pregnant women are aware of trustworthy and up-to-date information about the risks and benefits of vaccines and medications; take-up of vaccination is more likely in women who have the correct knowledge about the minimal risks involved (Lyerly et al., 2012). Care needs to be taken to ensure this group have realistic risk perceptions and are likely to adopt the recommended behaviours. Concerns about risk to their baby were common, so education about the benefits and risks to unborn babies are important. The willingness of many of Lynch et al.’s (2012) participants to change their views during focus groups highlights a critical role for education.

Participants reported disrupted routines and changes to relationships with others, including spouses, due to social distancing. This is of concern as it is well-known that social support is essential in enhancing resilience during times of crisis (Brooks et al., 2020a) while poor social support is associated with negative psychological outcomes (Goldmann & Galea, 2014), as is the isolation felt by people who are alone during quarantine in pandemics (Brooks et al., 2020b). One study in this review explored the relationship between social support and anxiety in pregnant women and found that social support was protective (Lee et al., 2006). Interestingly, this study also found that women who were pregnant during the SARS outbreak reported significantly higher affectionate support, positive social interaction and informational support than the pre-SARS cohort, although the groups did not differ on tangible support. However, participants in other studies reported feeling isolated and lonely. This raises the question of how to ensure that social support is maintained during times of social distancing – which is of course essential for the population as a whole, not just for vulnerable groups. Mental health campaigns aimed at encouraging communication via phone or internet during physical isolation may be useful. There is evidence that support from others with similar experiences can be particularly helpful to alleviate stress in pregnancy (McLeish & Redshaw, 2017) and that social media is a substantial source of support for pregnant women and new mothers (Baker & Yang, 2018); therefore, online or text message support groups specifically for pregnant women to support each other and discuss their concerns, fears and experiences may be beneficial.

Financial and occupational concerns were common, which is unsurprising as these have been cited as major stressors for anyone off work during pandemics (Brooks et al., 2020b). Pregnant women – and indeed the general public as a whole – would therefore benefit from ensuring they are aware of what financial assistance is available during the pandemic and how and when it can be claimed (HM Treasury, 2020). In particular, the COVID-19 outbreak may be stressful for pregnant women who are ‘critical workers’ and therefore expected to continue working (Cabinet Office, 2020) despite also being told they are a vulnerable group who should be “particularly stringent in following social distancing measures” (Public Health England, 2020). Organisations should help as much as possible, for example changing the work roles of pregnant women so they can work from home or away from the public where possible.

Participants in one study reported an overwhelming disruption in their expectations of birth, prenatal care and postnatal care, causing them to change their plans and preferences for birth. Additionally, maternity staff levels may be lower than usual during a pandemic due to reassignment of staff to other areas of the hospital or staff minimising contact with patients for their own protection. This raises the question of what is an acceptable level of care to provide to uninfected pregnant women during a pandemic and what could be considered neglectful (Dodgson et al., 2010). Guidance for healthcare professionals needs to be clear about which routine visits could be done over the phone or cancelled altogether, as well as how to provide appropriate care without exposing healthy women and newborns to illness. A solution may be designating a location and staff specifically for the care of healthy pregnant women (Rasmussen et al., 2008).

The reviewed literature showed that pregnant women often cope with concerns by taking drastic non-pharmaceutical precautions to avoid infection, which may affect all areas of their lives. Pregnant women may become hyper-vigilant with regard to monitoring the most current self-protection information available, hygiene practices and reducing contact with others. Of course, these practices are recommended in infectious disease outbreaks generally, and in themselves can be positive behaviours as they reduce the risk of infection. However, it is possible that such measures could also cause distress. More research is needed to explore the benefits and risks to mental health of prolonged hyper-vigilance, both for the population generally and during pregnancy and whether such behaviours predispose someone to post-outbreak mental ill-health.

Overall, this review highlights the fact that pregnant women are a highly vulnerable group in terms of psychological consequences during a pandemic; they need to care for both their own health and that of their unborn child, referred to as a ‘doubling of health responsibilities’ by Lohm et al. (2014). Planning for future pandemics should therefore make considerations specific to pregnant women to ensure their specific needs are addressed. Involving pregnant women in pandemic preparedness exercises would ensure that their voices are heard and specific concerns they may have are addressed, helping policy-makers to identify any gaps related to prenatal and postnatal care in current pandemic planning.

### Limitations

Data screening, extraction and analysis were carried out by only one author, due to the rapid nature of the review; in typical systematic reviews, it is preferable for double-screening to take place and for multiple reviewers to analyse the data to enhance the validity of the review. However, the resultant data was discussed with all authors as the paper went through multiple revisions prior to submission. Searches were limited to English-language papers due to lack of time to get foreign-language papers translated, meaning evidence may have been missed. The generalisability of the studies reviewed is not clear, as much may depend on the cultural context.

No standardised quality appraisal of the included papers was carried out, as is common in rapid evidence reviews (Haby et al., 2016). However, there were some particularly notable limitations to the literature, such as low response rates and a lack of quantitative research. It must also be noted that only one study (Lee et al., 2006) compared mental health outcomes for women pregnant during an outbreak with a pre-outbreak control group of pregnant women. For that reason, it is difficult to fully ascertain the mental health-related differences in being pregnant during a disease outbreak and being pregnant at any other time. There was also no research directly comparing pregnant women with the non-pregnant population during an outbreak, so again, we cannot say whether pregnant women are more likely to experience stress during an outbreak than the rest of the population.

### Conclusion

Pregnant women have specific needs during a pandemic and may be at risk of adverse psychological effects of the COVID-19 outbreak. This is important as there is a clear link between poor mental health in pregnant women and complications with the pregnancy. It is thus vital that they are well-informed about public health recommendations, which should include detailed description of benefits or lack of risk to unborn babies as well as a clear rationale for why prophylaxis or treatment is necessary. Virtual support groups (e.g. online) specifically for pregnant women to support each other may be useful although these should be informed by factual information and should not fuel anxiety. Healthcare professionals involved in the care of pregnant women should be aware of the most current guidance and ensure that they closely monitor mental health during pregnancy and where necessary provide early evidence-based care.

## Data Availability

Review article - no primary data.

## Disclaimer

The research was funded by the National Institute for Health Research Health Protection Research Unit (NIHR HPRU) in Emergency Preparedness and Response at King’s College London in partnership with Public Health England (PHE), in collaboration with the University of East Anglia. The views expressed are those of the author(s) and not necessarily those of the NHS, the NIHR, the Department of Health and Social Care or Public Health England.

## References

American College of Obstetricians and Gynecologists. (2017). Hospital disaster preparedness for obstetricians and facilities providing maternity care. Obstetrics and Gynecology, 130(6), e291–e297.

Asghari, E., Faramarzi, M. & Mohammmadi, A.K. (2016). The effect of cognitive behavioural therapy on anxiety, depression and stress in women with preeclampsia. Journal of Clinical & Diagnostic Research, 10(11), QC04–1C07.

Baker, B. & Yang, I. Social media as social support in pregnancy and the postpartum. Sexual & Reproductive Healthcare, 17, 31–34.

Bessett, D. (2010). Negotiating normalization: The perils of producing pregnancy symptoms in prenatal care. Social Science & Medicine, 71, 370–377.

Braun, V. & Clarke, V. (2006). Using thematic analysis in psychology. Qualitative Research in Psychology, 3(2), 77–101.

Brooks, S.K., Amlot, R., Rubin, G.J. & Greenberg, N. (2020a). Psychological resilience and post-traumatic growth in disaster-exposed organisations: Overview of the literature. BMJ Military Health, 166(1), 52–56.

Brooks, S.K., Webster, R.K., Smith, L.E., Woodland, L., Wessely, S., Greenberg, N. & Rubin, G.J. (2020b). The psychological impact of quarantine and how to reduce it: rapid review of the evidence. The Lancet, 395(10227), 912–920.

Cabinet Office. (2020). Guidance for schools, childcare providers, colleges and local authorities in England on maintaining educational provision. Available online: https://www.gov.uk/government/publications/coronavirus-covid-19-maintaining-educational-provision/guidance-for-schools-colleges-and-local-authorities-on-maintaining-educational-provision [accessed 12 April 2020]

Chakhtoura, N., Hazra, R. & Spong, C.Y. (2019). Zika virus: A public health perspective. Current Opinion in Obstetrics and Gynecology, 30(2), 116–122.

Dashraath, P., Jing Lin Jeslyn, W., Mei Xian Karen, L., Li Min, L., Sarah, L., Biswas, A., Arjandas Choolani, M., Mattar, C. & Lin, S.L. (2020). Coronavirus disease 2019 (COVID-19) pandemic and pregnancy. American Journal of Obstetrics and Gynecology, doi: 10.1016/j.ajog.2020.03.021.

Dodgson, J.E., Tarrant, M., Chee, Y.O. & Watkins, A. (2010). New mothers’ experiences of social disruption and isolation during the severe acute respiratory syndrome outbreak in Hong Kong. Nursing & Health Sciences, 12(2), 198–204.

Fridman, D., Steinberg, E., Azhar, E., Weedon, J. & Wilson, T.E. (2011). Predicators of H1N1 vaccination in pregnancy. American Journal of Obstetrics and Gynecology, 204(6, S1), S124–127.

Gardner, P.J. & Moallef, P. (2015). Psychological impact on SARS survivors: Critical review of the English language literature. Canadian Psychology, 56(1), 123–135.

Goldmann, E. & Galea, S. (2014). Mental health consequences of disasters. Annual Review of Public Health, 35, 169–183.

Goodall, C., Sabo, J., Cline, R. & Egbert, N. (2012). Threat, efficacy, and uncertainty in the first 5 months of national print and electronic news coverage of the H1N1 virus. Journal of Health Communication, 17(3), 338–355.

Goodman, J.H., Guarino, A., Chenausky, K., Klein, L., Prager, J., Petersen, R., Forget, A. & Freeman, M. (2014). CALM pregnancy: results of a pilot study of mindfulness-based cognitive therapy for perinatal anxiety. Archives of Women’s Mental Health, 17(5), 373–387.

Grupe, D.W. & Nitschke, J.B. (2013). Uncertainty and anticipation in anxiety: An integrated neurobiological and psychological perspective. Nature Reviews Neuroscience, 14(7), 488–501.

Haby, M.M., Chapman, E., Clark, R., Barreto, J., Reveiz, L. & Lavis, J.N. (2016). What are the best methodologies for rapid reviews of the research evidence for evidence-informed decision making in health policy and practice: a rapid review. Health Research Policy and Systems, 14(1), 83.

HM Treasury. (2020). Support for those affected by COVID-19. Available online: https://www.gov.uk/government/publications/support-for-those-affected-by-covid-19/support-for-those-affected-by-covid-19 [accessed 12 April 2020]

Jamieson, D.J., Ellis, J.E., Jernigan, D.B. & Treadwell, T.A. (2006b). Emerging infectious disease outbreaks: Old lessons and new challenges for obstetrician-gynecologists. American Journal of Obstetrics and Gynecology, 194, 1546–1555.

Jamieson, D.J., Theiler, R. & Rasmussen, S.A. (2006a). Emerging infections and pregnancy. Emerging I nfectious Diseases, 12(11), 1638–1643.

Jarvis, S. (2020). COVID-19: What you need to know about coronavirus and pregnancy. Available online: https://patient.info/news-and-features/covid-19-what-you-need-to-know-about-coronavirus-and-pregnancy [accessed 03 April 2020]

Kahn, M. & Cristoferi, C. (2020). Anxiety, anger and hope as women face childbirth during coronavirus pandemic. Available online: https://uk.reuters.com/article/us-health-coronavirus-europe-childbirth/anxiety-anger-and-hope-as-women-face-childbirth-during-coronavirus-pandemic-idUKKBN21E1O2 [accessed 12 April 2020]

Lam, C.M., Wong, S.F., Leung, T.N., Chow, K.M., Yu, W.C., Wong, T.Y., Lai, S.T. & Ho, L.C. (2004). A case-controlled study comparing clinical course and outcomes of pregnant and non-pregnant women with severe acute respiratory syndrome. BJOG: an International Journal of Obstetrics and Gynaecology, 111, 771–774.

Lee, C-H., Huang, N., Chang, H-J., Hsu, Y-J., Wang, M.C. & Chou, Y-J. (2005). The immediate effects of the severe acute respiratory syndrome (SARS) epidemic on childbirth in Taiwan. BMC P ublic Health, 5, 30.

Lee, D.T., Sahota, D., Leung, T.N., et al. (2006). Psychological responses of pregnant women to an infectious outbreak: A case-control study of the 2003 SARS outbreak in Hong Kong. Journal of Psychosomatic Research, 61(5), 707–713.

Linde, A.R. & Siqueira, C.E. (2018). Women’s lives in times of Zika: mosquito-controlled lives? Cadernos de saude publica, 34(5), e00178917.

Lohm, D., Flowers, P., Stephenson, N., Waller, E. & Davis, M.D.M. (2014). Biography, pandemic time and risk: Pregnant women reflecting on their experiences of the 2009 influenza pandemic. Health, 18(5), 493–508.

Lyerly, A.D., Namey, E.E., Gray, B., Swamy, G. & Faden, R.R. (2012). Women’s views about participating in research while pregnant. IRB: Ethics & Human Research, 34(4), 1–8.

Lynch, M.M., Mitchell, E.W., Williams, J.L., Brumbaugh, K., Jones-Bell, M., Pinkney, D.E., Layton, C.M., Mersereau, P.W., Kendrick, J.S., Eguino Medina, P. & Rojas Smith, L. (2012). Pregnant and recently pregnant women’s perceptions about influenza A pandemic (H1N1) 2009: Implications for public health and provider communication. Maternal and Child Health Journal, 16, 1657–1664.

Maher, M.J. (2019). Emergency preparedness in obstetrics: Meeting unexpected key challenges. Journal of Perinatal & Neonatal Nursing, 33(3), 238–245.

McLeish, J. & Redshaw, M. (2017). Mothers’ accounts of the impact on emotional wellbeing of organised peer support in pregnancy and early parenthood: a qualitative study. BMC Pregnancy and Childbirth, 17, 28.

Meireles, J.F.F., Neves, C.M., da Rocha Morgado, F.F. & Ferreira, M.E.C. (2017). Zika vírus and pregnant women: A psychological approach. Psychology & Health, 32(7), 798–809.

Melander, H. (2002). Experiences of fears associated with pregnancy and childbirth: A study of 329 pregnant women. Birth, 29(2), 101–111.

Mosby, L.G., Rasmussen, S.A. & Jamieson, D.J. (2011). 2009 pandemic influenza A (H1H1) in pregnancy: a systematic review of the literature. American Journal of Obstetrics and Gynecology, 205(1), 10–18.

Ng, J., Sham, A., Leng Tang, P. & Fung, S. (2004). SARS: Pregnant women’s fears and perceptions. British Journal of Midwifery, 12(11), 698–703.

Oppenheim, M. (2020). Pregnant women forced to give birth without support in hospital amid coronavirus outbreak. The Independent. Available online: https://www.independent.co.uk/news/uk/home-news/coronavirus-pregnant-women-birth-hospital-nhs-parents-advice-a9439391.html [accessed 05 April 2020]

Ozer, A., Arikan, D.C., Kirecci, E. & Ekerbicer, H.C. (2010). Status of pandemic influenza vaccination and factors affecting it in pregnant women in Kahramanmaras, an Eastern Mediterranean city of Turkey. PLoS ONE, 5(12), e14177.

Pais, M. & Pai, M.V. (2018). Stress among pregnant women: A systematic review. Journal o f Clinical and Diagnostic Research, 12(5), LE01–LE04.

Patterson, J., Hollins Martin, C. & Karatzias, T. (2018). PTSD post-childbirth: A systematic review of women’s and midwives’ subjective experiences of care provider interaction. Journal of Reproductive and Infant Psychology, 37(1), 56–83.

Perrin, P.C., McCabe, L. & Everly Jr, G.S. (2009). Preparing for an influenza pandemic: Mental health considerations. Prehospital a nd Disaster Medicine, 24(3), 223–230.

Public Health England. (2020). Guidance on social distancing for everyone in the UK. Available online: https://www.gov.uk/government/publications/covid-19-guidance-on-social-distancing-and-for-vulnerable-people/guidance-on-social-distancing-for-everyone-in-the-uk-and-protecting-older-people-and-vulnerable-adults [accessed 10 April 2020]

Rasmussen, S.A., Jamieson, D.J. & Bresee, J.S. (2008). Pandemic influenza and pregnant women. Emerging Infectious Diseases, 14(1), 95–100.

Rawlinson, K. (2020). Coronavirus latest: April 3, at a glance. The Guardian. Available online: https://www.theguardian.com/world/2020/apr/04/coronavirus-latest-at-a-glance [accessed 12 April 2020]

Sakaguchi, S., Weitzner, B., Carey, N., Bozzo, P., Mirdamadi, K., Samuel, N., Koren, G. & Einarson, A. (2011). Pregnant women’s perception of risk with use of the H1N1 vaccine. Journal o f Obstetrics and Gynaecology Canada, 33(5), 460–467.

Sasaki, T-K., Yoshida, A. & Kotake, K. (2013). Attitudes about the 2009 H1N1 influenza pandemic among pregnant Japanese women and the use of the Japanese municipality as a source of information. Southeast Asian Journal of Tropical Medicine and Public Health, 44(3), 388–399.

Schmid, P., Rauber, D., Betsch, C., Lidolt, G. & Denker, M-L. (2017). Barriers of influenza vaccination intention and behavior – A systematic review of influenza vaccine hesitancy, 2005-2016. PLoS ONE, 12(1), e0170550.

Schwartz, D.A. & Graham, A.L. (2020). Potential maternal and infant outcomes from (Wuhan) Coronavirus 2019-nCoV infecting pregnant women: Lessons from SARS, MERS, and other human coronavirus infections. Viruses, 12(2), 194.

Sim, J.A., Ulanika, A.A., Katikireddi, S.V. & Gorman, D. (2011). ‘Out of two bad choices, I took the slightly better one’: Vaccination dilemmas for Scottish and Polish migrant women during the H1N1 influenza pandemic. Public Health, 125(8), 505–511.

Steelfisher, G.K., Blendon, R.J., Bekheit, M.M., Mitchell, E.W., Williams, J., Lubell, K., Peugh, J. & DiSogra, C.A. (2011). Novel pandemic A (H1N1) influenza vaccination among pregnant women: motivators and barriers. American Journal of Obstetrics & Gynecology, 204(6 Suppl 1), S116–123.

Stevenson, A. (2020). ‘I felt like crying’: Coronavirus shakes China’s expecting mothers. The New York Times. Available online: https://www.nytimes.com/2020/02/25/business/coronavirus-china-pregnant.html [accessed 12 April 2020]

Williamson, V., Murphy, D. & Greenberg, N. (2020). COVID-19 and experiences of moral injury in front-line key workers. Occupational Medicine, https://doi.org/10.1093/occmed/kqaa052

World Health Organization. (2010). Pregnancy and pandemic influenza A (H1N1) 2009: Information for programme managers and clinicians. Available online: https://www.ncbi.nlm.nih.gov/books/NBK200794/pdf/Bookshelf_NBK200794.pdf [accessed 02 April 2020]

World Health Organization. (2014). A brief guide to emerging infectious diseases and zoonoses. WHO Regional Office for South-East Asia. Available online: https://apps.who.int/iris/handle/10665/204722 [accessed 12 April 2020]

World Health Organization. (2017). Rapid reviews to strengthen health policy and systems: a practical guide. Available online: https://www.who.int/alliance-hpsr/resources/publications/rapid-review-guide/en/ [accessed 05 April 2020]

Yuen, C.Y.S. & Tarrant, M. Determinants of uptake of influenza vaccination among pregnant women – A systematic review. Vaccine, 32, 4602–4613.

